# Local immunoglobulin expression in myositis is associated with interferon gamma signaling and correlates with disease activity

**DOI:** 10.1101/2025.06.03.25328909

**Authors:** Iago Pinal-Fernandez, Caroline Metz, Maria Casal-Dominguez, Katherine Pak, Raphael Kirou, Stefania Dell’Orso, Faiza Naz, Shamima Islam, Gustavo Gutierrez-Cruz, Werner Stenzel, Albert Selva-O’Callaghan, Jose C. Milisenda, Andrew L. Mammen

**Author notes:** Address correspondence to: Andrew L. Mammen, M.D., Ph.D., or Iago Pinal-Fernandez, M.D., Ph.D., Ph.D., Muscle Disease Section, National Institute of Arthritis and Musculoskeletal and Skin Diseases, National Institutes of Health, 50 South Drive, Room 1141, Building 50, MSC 8024, Bethesda, MD 20892. or. These authors contributed equally to this project (Iago Pinal-Fernandez, Caroline Metz). **Competing interests**: None. **Contributorship:** All authors contributed to the development of the manuscript, including interpretation of results, substantive review of drafts, and approval of the final draft for submission. IPF and ALM are the guarantors. **Funding**: This study was funded, in part, by the Intramural Research Program of the National Institute of Arthritis and Musculoskeletal and Skin Diseases, National Institutes of Health. **Ethical approval information:** All biopsies were from subjects enrolled in institutional review board (IRB)-approved longitudinal cohorts in the National Institutes of Health, the Johns Hopkins, the Clinic Hospital, the Vall d’Hebron Hospital, and the Charité-Universitätsmedizin Berlin. **Patient and public involvement**: Patients and/or the public were not involved in the design, conduct, reporting, or dissemination plans of this research. **Data sharing statement:** Any anonymized data not published within the article will be shared by request from any qualified investigator.

## Abstract

**Objectives:** Autoantibodies may play a role in the pathogenesis of myositis and are locally produced within muscle tissue. This study aimed to characterize the local expression of immunoglobulin genes across different subgroups of myositis, identify pathways associated with this expression, and evaluate correlations with disease activity.

**Methods:** Bulk RNA sequencing was performed on muscle biopsies from 289 individuals, including patients with various forms of myositis and healthy controls. Expression levels of immunoglobulin gene regions were compared across clinical and autoantibody-defined subgroups. Pathway enrichment analysis and unsupervised clustering were conducted, and correlations between immunoglobulin gene expression and disease activity were assessed.

**Results:** Local immunoglobulin gene expression was highest in inclusion body myositis (IBM) and antisynthetase syndrome (ASyS), followed by dermatomyositis, and lowest in immune-mediated necrotizing myopathy. Among isotypes, IgG, IgA, and IgM predominated, while IgD and IgE expression was minimal. Certain immunoglobulin VJ segments were more frequently used across all patients, with no significant differences in specific region usage between patient groups. Immunoglobulin gene expression strongly correlated with disease activity, particularly in patients with anti-Mi2, anti-MDA5, anti-Jo1 autoantibodies, and IBM. Pathway analysis revealed a robust association between immunoglobulin expression and interferon-gamma (IFN-γ) signaling. Unsupervised clustering based solely on immunoglobulin gene expression clearly separated healthy controls from patients with ASyS and IBM.

**Conclusions:** Immunoglobulin is locally expressed in the muscle of patients with myositis, particularly IBM and ASyS. This expression correlates with disease activity, involves preferential usage of specific isotypes and gene segments, and is closely linked to IFN-γ-associated immune pathways.

## INTRODUCTION

Inflammatory myopathies are a heterogeneous group of autoimmune disorders characterized by chronic muscle inflammation and varying degrees of extramuscular involvement, most commonly affecting the skin, lungs, and joints (1). The major subtypes include dermatomyositis (DM), antisynthetase syndrome (ASyS), inclusion body myositis (IBM), and immune-mediated necrotizing myopathy (IMNM) (1). A defining feature of these conditions is the presence of myositis-specific autoantibodies, which delineate clinically and mechanistically distinct subgroups and are associated with differences in pathogenesis, histopathology, disease trajectory, and treatment response (1, 2).

Accumulating evidence suggests that antibodies may be generated locally within inflamed muscle tissue, potentially explaining the selective involvement of muscle tissue and certain muscle groups in these diseases. Histopathological analyses of muscle biopsies from patients with myositis have identified antigen-experienced B cells and plasma cells infiltrating the tissue. In particular, biopsies from individuals IBM show abundant CD138⁺ plasma cells, and transcriptomic studies reveal that immunoglobulin (Ig) genes are among the most upregulated in affected muscle samples (3–5). Laser-capture microdissection followed by Ig transcript sequencing has demonstrated clonally related B cells and plasma cells dispersed across myositis lesions, further supporting the concept of local B-cell maturation and differentiation into antibody-secreting cells (6). In DM and overlap myositis, muscle biopsies can exhibit lymphoid follicles and B-cell aggregates resembling germinal centers, which may serve as niches for in situ antibody production (7, 8). Similarly, in ASyS, muscle biopsies frequently display perimysial clusters of CD20⁺ B cells and CD138⁺ plasma cells (9). Collectively, these findings suggest that inflamed muscle in myositis provides a permissive microenvironment for B-cell activation, maturation, and local immunoglobulin synthesis (3, 4, 6).

To date, no large-scale transcriptomic study has systematically quantified immunoglobulin gene expression across the full spectrum of myositis subtypes or explored its relationship with clinical disease activity and associated inflammatory pathways. In this study, we address this knowledge gap by analyzing RNA-sequencing data from muscle biopsies of patients with DM, ASyS, IMNM, and IBM. We comprehensively profile the expression of immunoglobulin heavy- and light-chain regions in each subtype, assess their association with clinical markers of disease severity, and examine their relationship with key inflammatory signatures.

## METHODS

### Patients

Muscle biopsies from myopathy patients were collected from Institutional Review Board (IRB)-approved longitudinal cohorts at the National Institutes of Health (Bethesda, MD), Johns Hopkins Myositis Center (Baltimore, MD), Vall d’Hebron Hospital (Barcelona), Clinic Hospital (Barcelona), and Charité-Universitätsmedizin (Berlin). Patients were classified as IBM if they met Lloyd’s criteria (10). Autoantibodies were detected by ELISA, immunoprecipitation of *in vitro* transcription and translation-generated proteins (IVTT-IP), EUROLINE myositis profile line blotting, or immunoprecipitation from ^35^S-methionine-labeled cell lysates. Patients testing positive for a myositis-specific autoantibody were classified based on their myositis-specific autoantibody according to the NIH criteria (11): those positive for anti-Mi2, anti-NXP2, anti-MDA5, or anti-TIF1γ were classified as DM; those positive for anti-Jo1, were classified as ASyS; and those with autoantibodies against SRP or HMGCR were classified as IMNM. Patients with less prevalent autoantibodies were excluded from the analysis.

### RNA sequencing

Bulk RNAseq was performed on frozen muscle biopsy specimens as previously described (4, 12–16). Briefly, muscle biopsies underwent immediate flash freezing and were stored at -80°C across all contributing centers. Samples were then transported in dry ice to the NIH and processed uniformly to prepare the library and conduct the analysis. RNA was extracted with TRIzol. Libraries were either prepared with the NeoPrep system according to the TruSeq Stranded mRNA Library Prep protocol (Illumina, San Diego, CA) or with the NEBNext Poly(A) mRNA Magnetic Isolation Module and Ultra^™^ II Directional RNA Library Prep Kit for Illumina (New England BioLabs, ref. #E7490, and #E7760).

### Statistical and bioinformatic analysis

For RNAseq analysis, sequencing reads were demultiplexed using bcl2fastq/2.20.0 and preprocessed using fastp/0.21.0. The abundance of each gene was determined using Salmon/1.5.2. Counts were normalized using the Trimmed Means of M values (TMM) from edgeR/3.34.1 for graphical analysis. Differential expression was performed using limma/3.48.3. Pathway enrichment was assessed using ReactomePA (v1.48.0) to perform gene set enrichment analysis (GSEA) against the Reactome database, and results were visualized with enrichplot (v1.24.4). Gene lists were curated from the HUGO Gene Nomenclature Committee (HGNC). Relative expression was expressed as the log2 fold-change (logFC). Where applicable, P-values were adjusted for multiple comparisons using the Benjamini–Hochberg procedure, with a false discovery rate threshold of q < 0.05 considered statistically significant.

## RESULTS

Muscle biopsies from 289 individuals were analyzed, comprising 252 patients with inflammatory myopathies and 37 healthy controls. Patients were categorized into clinically and serologically defined subgroups: DM (n=82, including anti-Mi2 [n=22], anti-NXP2 [n=21], anti-TIF1γ [n=28], and anti-MDA5 [n=11]), ASyS with anti-Jo1 antibodies (n=37), IMNM (n=80, including anti-HMGCR [n=60] and anti-SRP [n=20]), and IBM (n=53) (Supplementary Table 1).

### Elevated Local Immunoglobulin Expression in IBM and Antisynthetase Syndrome

IgG, IgM, and IgA were the predominant isotypes detected, while overexpression of IgD and IgE was negligible across all groups. Expression levels of IgG, IgM, and IgA were strongly correlated with one another, while their associations with IgD and IgE were comparatively weak (Figure 2).

**Figure 1.**
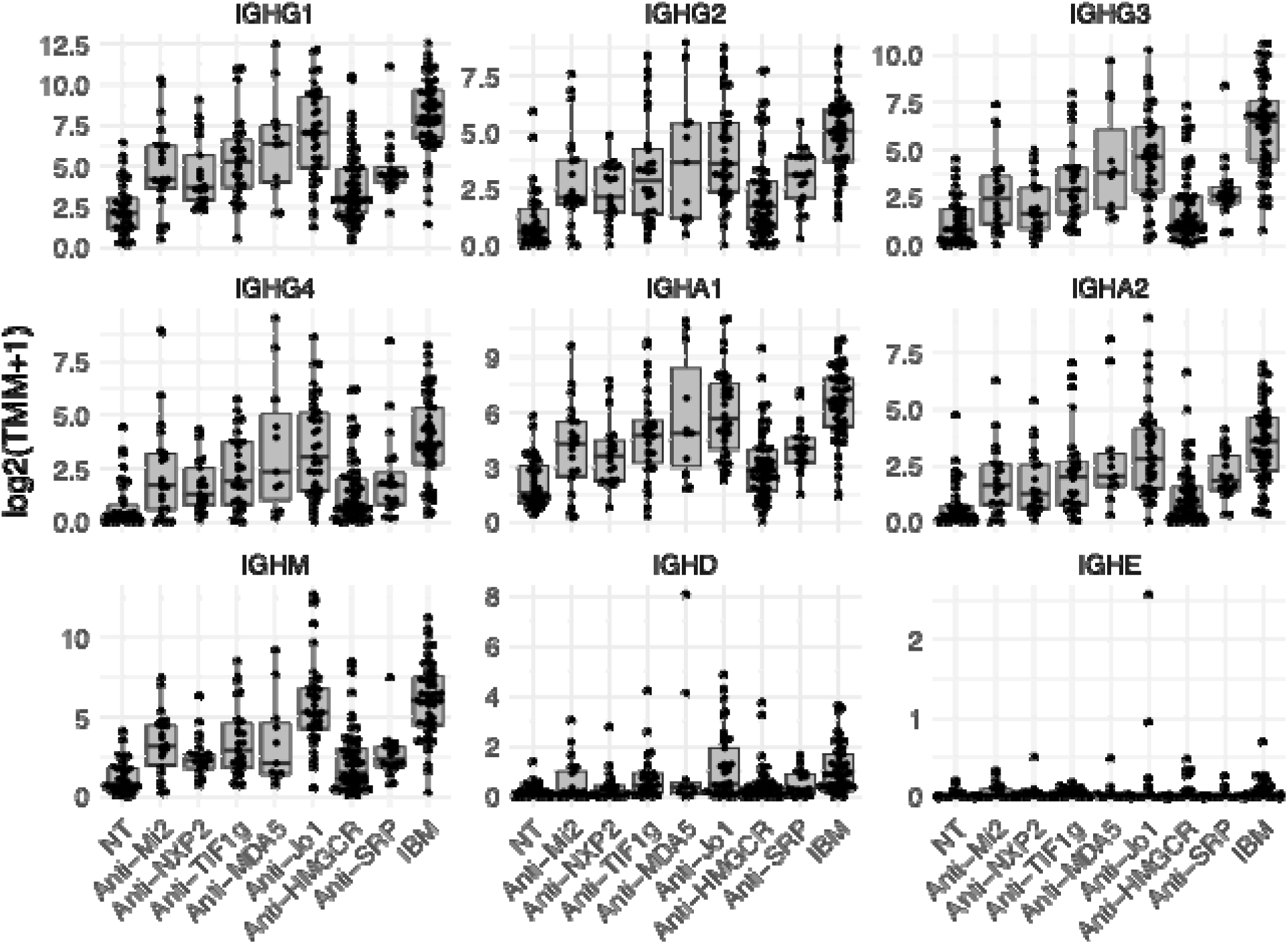
Expression of immunoglobulin heavy chain constant region genes across disease groups. Each point represents an individual sample. Expression is presented as trimmed mean of M values (TMM). Normal muscle biopsy: NT; inclusion body myositis: IBM.

**Figure 2.**
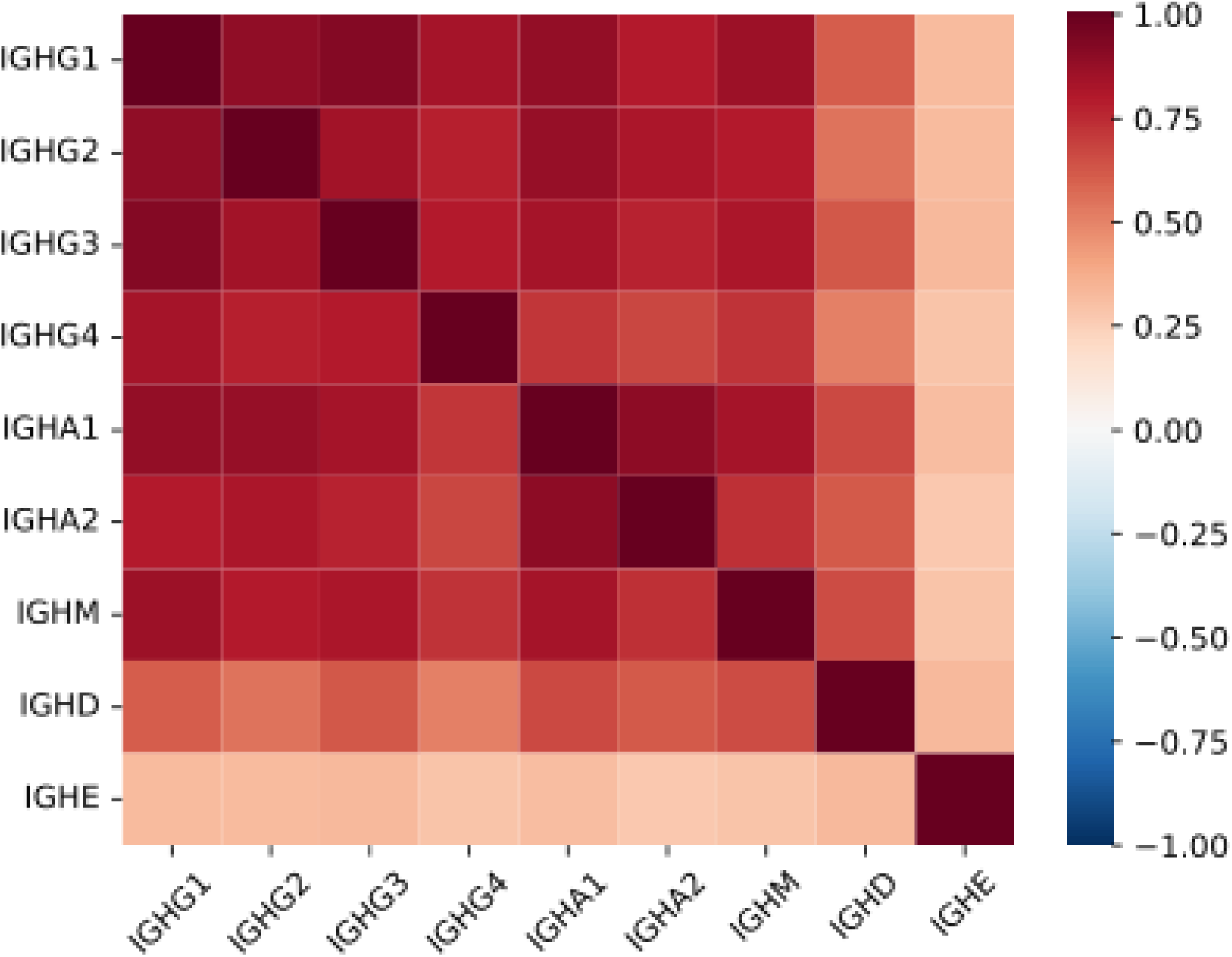
Spearman correlation matrix of immunoglobulin heavy chain genes.

Muscle biopsies from patients with IBM and anti-Jo1 ASyS exhibited significantly higher expression of immunoglobulin genes compared to other subgroups (Table 1, Figure 1). In IBM, multiple immunoglobulin heavy-chain constant regions were markedly upregulated, particularly those encoding IgG isotypes. For example, the constant region of IgG3 showed a log fold change (logFC) of 4.2 (q = 3.99 × 10⁻² ), while that of IgG1 had a logFC of 3.7 (q = 4.42 × 10⁻²¹) compared to other muscle biopsies. Alternatively, anti-Jo1 samples demonstrated relatively elevated expression of IgM and IgA constant regions (IGHM logFC = 3.1, q = 4.43 × 10⁻² ; IGHA1 logFC = 2.0, q = 3.59 × 10⁻ ). DM biopsies showed intermediate immunoglobulin expression, while IMNM had the lowest levels (Table 1, Figure 1).

**Table 1.** Expression of immunoglobulin heavy chain constant region genes across disease groups, shown as comparisons versus all other muscle biopsies (vs. ALL) and versus normal muscle biopsies (vs. NT) . DM: dermatomyositis; IMNM: immune-mediated necrotizing myopathy; IBM: inclusion body myositis; NT: normal muscle biopsy.

| DM vs. ALL |  |  | Mi2 vs. ALL |  |  | NXP2 vs. ALL |  |  | MDA5 vs. ALL |  |  | TIF1γvs. ALL |  |  |
| --- | --- | --- | --- | --- | --- | --- | --- | --- | --- | --- | --- | --- | --- | --- |
| Gene | LogFC | q-val | Gene | LogFC | q-val | Gene | LogFC | q-val | Gene | LogFC | q-val | Gene | LogFC | q-val |
| IGHM | -0.4 | 0.450 | IGHD | -0.9 | 0.304 | IGHG3 | -1.4 | 0.385 | IGHA2 | 1.5 | 0.596 | IGHD | 0.4 | 0.652 |
| IGHG3 | -0.4 | 0.459 | IGHG2 | -0.7 | 0.559 | IGHM | -1.2 | 0.436 | IGHG4 | 1.6 | 0.615 | IGHG2 | 0.4 | 0.714 |
| IGHE | 0.2 | 0.534 | IGHA1 | -0.5 | 0.596 | IGHA1 | -0.8 | 0.522 | IGHA1 | 1.3 | 0.652 | IGHA1 | 0.4 | 0.727 |
| IGHG1 | -0.2 | 0.722 | IGHG3 | -0.6 | 0.598 | IGHG1 | -0.9 | 0.550 | IGHG3 | 1.3 | 0.687 | IGHE | 0.2 | 0.745 |
| IGHA2 | 0.2 | 0.742 | IGHG1 | -0.5 | 0.692 | IGHG2 | -0.9 | 0.574 | IGHD | 1.1 | 0.700 | IGHA2 | 0.3 | 0.809 |
| IGHG2 | -0.2 | 0.761 | IGHG4 | -0.5 | 0.706 | IGHD | -0.6 | 0.644 | IGHG1 | 1.2 | 0.706 | IGHG1 | 0.2 | 0.906 |
| IGHD | -0.1 | 0.766 | IGHA2 | -0.2 | 0.837 | IGHG4 | -0.4 | 0.817 | IGHG2 | 1.0 | 0.719 | IGHG3 | 0.1 | 0.930 |
| IGHA1 | -0.1 | 0.848 | IGHM | 0.0 | 0.993 | IGHA2 | -0.3 | 0.821 | IGHE | 0.2 | 0.888 | IGHG4 | -0.1 | 0.957 |
| IGHG4 | -0.1 | 0.922 | IGHE | 0.0 | 0.999 | IGHE | 0.1 | 0.869 | IGHM | -0.1 | 0.984 | IGHM | 0.0 | 0.978 |

| JO1 vs. ALL |  |  | IMNM vs. ALL |  |  | HMGR vs. ALL |  |  | SRP vs. ALL |  |  | IBM vs. ALL |  |  |
| --- | --- | --- | --- | --- | --- | --- | --- | --- | --- | --- | --- | --- | --- | --- |
| Gene | LogFC | q-val | Gene | LogFC | q-val | Gene | LogFC | q-val | Gene | LogFC | q-val | Gene | LogFC | q-val |
| IGHM | 3.1 | 4.43e-09 | IGHM | -2.1 | 0.001 | IGHA2 | -2.2 | 0.002 | IGHM | -1.0 | 0.996 | IGHG3 | 4.2 | 3.99e-25 |
| IGHA1 | 2.0 | 3.59e-04 | IGHA1 | -1.6 | 0.002 | IGHA1 | -1.9 | 0.004 | IGHE | -0.3 | 0.996 | IGHG1 | 3.7 | 4.42e-21 |
| IGHA2 | 2.0 | 0.001 | IGHA2 | -1.7 | 0.004 | IGHM | -2.1 | 0.006 | IGHA2 | 0.4 | 0.996 | IGHM | 3.6 | 2.03e-19 |
| IGHG1 | 2.1 | 0.001 | IGHG1 | -1.8 | 0.005 | IGHG1 | -2.0 | 0.009 | IGHG1 | -0.4 | 0.996 | IGHA1 | 2.9 | 1.09e-14 |
| IGHG4 | 2.1 | 0.004 | IGHG3 | -1.8 | 0.006 | IGHG3 | -2.1 | 0.010 | IGHD | -0.3 | 0.996 | IGHG2 | 3.0 | 2.85e-13 |
| IGHG3 | 2.0 | 0.008 | IGHD | -1.1 | 0.038 | IGHG2 | -1.6 | 0.043 | IGHG3 | -0.3 | 0.996 | IGHA2 | 2.8 | 5.15e-12 |
| IGHG2 | 1.7 | 0.022 | IGHG4 | -1.4 | 0.042 | IGHG4 | -1.7 | 0.050 | IGHA1 | -0.3 | 0.996 | IGHG4 | 2.9 | 4.43e-11 |
| IGHD | 1.3 | 0.092 | IGHG2 | -1.2 | 0.056 | IGHD | -1.2 | 0.052 | IGHG2 | 0.2 | 0.996 | IGHD | 1.9 | 2.29e-06 |
| IGHE | -0.1 | 0.987 | IGHE | -0.4 | 0.165 | IGHE | -0.4 | 0.318 | IGHG4 | -0.1 | 0.996 | IGHE | 0.5 | 0.123 |

| DM vs. NT |  |  | Mi2 vs. NT |  |  | NXP2 vs. NT |  |  | MDA5 vs. NT |  |  | TIF1γvs. NT |  |  |
| --- | --- | --- | --- | --- | --- | --- | --- | --- | --- | --- | --- | --- | --- | --- |
| Gene | LogFC | q-val | Gene | LogFC | q-val | Gene | LogFC | q-val | Gene | LogFC | q-val | Gene | LogFC | q-val |
| IGHM | 3.0 | 3.05e-05 | IGHM | 3.3 | 2.32e-05 | IGHM | 2.2 | 2.07e-04 | IGHG3 | 4.6 | 8.84e-05 | IGHM | 3.2 | 9.68e-05 |
| IGHA2 | 3.1 | 1.63e-04 | IGHG1 | 2.9 | 5.70e-04 | IGHG4 | 2.9 | 2.81e-04 | IGHA2 | 4.3 | 1.05e-04 | IGHG1 | 3.5 | 1.15e-04 |
| IGHG3 | 3.1 | 1.82e-04 | IGHA2 | 2.7 | 0.001 | IGHG1 | 2.5 | 6.63e-04 | IGHG1 | 4.4 | 1.08e-04 | IGHG3 | 3.5 | 1.80e-04 |
| IGHG1 | 3.3 | 1.96e-04 | IGHG3 | 2.8 | 0.002 | IGHA2 | 2.6 | 0.001 | IGHA1 | 3.8 | 4.72e-04 | IGHA1 | 2.9 | 6.20e-04 |
| IGHG4 | 3.2 | 0.001 | IGHA1 | 2.1 | 0.009 | IGHA1 | 1.8 | 0.003 | IGHG4 | 4.7 | 6.72e-04 | IGHG2 | 3.5 | 7.32e-04 |
| IGHG2 | 3.0 | 0.001 | IGHG4 | 2.8 | 0.021 | IGHG2 | 2.3 | 0.004 | IGHG2 | 4.1 | 8.31e-04 | IGHA2 | 3.2 | 0.001 |
| IGHA1 | 2.5 | 0.002 | IGHG2 | 2.5 | 0.026 | IGHG3 | 2.1 | 0.016 | IGHM | 3.2 | 0.002 | IGHG4 | 3.2 | 0.002 |
| IGHD | 1.4 | 0.027 | IGHD | 0.7 | 0.399 | IGHD | 1.0 | 0.190 | IGHD | 2.5 | 0.012 | IGHD | 1.9 | 0.004 |
| IGHE | 0.3 | 0.363 | IGHE | 0.1 | 0.889 | IGHE | 0.3 | 0.625 | IGHE | 0.3 | 0.636 | IGHE | 0.4 | 0.309 |

| JO1 vs. NT |  |  | IMNM vs. NT |  |  | HMGR vs. NT |  |  | SRP vs. NT |  |  | IBM vs. NT |  |  |
| --- | --- | --- | --- | --- | --- | --- | --- | --- | --- | --- | --- | --- | --- | --- |
| Gene | LogFC | q-val | Gene | LogFC | q-val | Gene | LogFC | q-val | Gene | LogFC | q-val | Gene | LogFC | q-val |
| IGHA1 | 4.3 | 3.92e-08 | IGHG1 | 2.1 | 0.002 | IGHG1 | 1.8 | 0.010 | IGHA2 | 3.3 | 4.92e-06 | IGHG2 | 5.7 | 4.16e-10 |
| IGHG1 | 5.2 | 2.20e-07 | IGHG3 | 2.0 | 0.011 | IGHG4 | 1.9 | 0.025 | IGHG1 | 2.9 | 1.04e-05 | IGHA1 | 5.0 | 2.71e-09 |
| IGHM | 5.9 | 5.07e-07 | IGHG4 | 2.2 | 0.011 | IGHM | 1.6 | 0.036 | IGHG2 | 3.3 | 1.25e-05 | IGHA2 | 5.3 | 1.47e-08 |
| IGHA2 | 4.7 | 6.28e-07 | IGHM | 1.8 | 0.014 | IGHG3 | 1.7 | 0.037 | IGHA1 | 2.3 | 1.41e-05 | IGHG1 | 6.5 | 2.91e-08 |
| IGHG3 | 5.0 | 4.35e-06 | IGHG2 | 2.2 | 0.016 | IGHG2 | 1.9 | 0.052 | IGHG3 | 3.0 | 1.18e-04 | IGHG4 | 5.8 | 1.36e-07 |
| IGHG2 | 4.6 | 7.62e-06 | IGHA1 | 1.4 | 0.027 | IGHA1 | 1.1 | 0.100 | IGHM | 2.3 | 1.96e-04 | IGHM | 6.3 | 2.24e-07 |
| IGHG4 | 5.1 | 8.86e-06 | IGHA2 | 1.7 | 0.043 | IGHA2 | 1.2 | 0.160 | IGHG4 | 3.2 | 0.001 | IGHD | 3.1 | 2.96e-07 |
| IGHD | 2.6 | 5.76e-04 | IGHD | 0.7 | 0.199 | IGHD | 0.6 | 0.336 | IGHD | 1.2 | 0.064 | IGHG3 | 6.9 | 5.15e-07 |
| IGHE | 0.1 | 0.904 | IGHE | -0.2 | 0.603 | IGHE | -0.2 | 0.636 | IGHE | 0.0 | 0.924 | IGHE | 0.6 | 0.072 |

### Local Immunoglobulin Expression Correlates with Disease Activity

Immunoglobulin gene expression was significantly positively correlated with multiple transcriptomic markers of disease activity, including type I interferon-inducible genes (ISG15, MX1), type II interferon-inducible genes (GBP2, IFI30), T-cell markers (CD3E, CD4, CD8), macrophage markers (CD14, CD68), and muscle regeneration markers (MYH3, MYH8, NCAM1, PAX7). In contrast, immunoglobulin expression was inversely associated with mitochondrial genes (MT-CO1, MT-CO2) and genes encoding mature muscle structural proteins (ACTA1, MYH1, MYH2). Across all patients, higher levels of intramuscular immunoglobulin transcripts were associated with increased disease activity (Figure 3).

**Figure 3.**
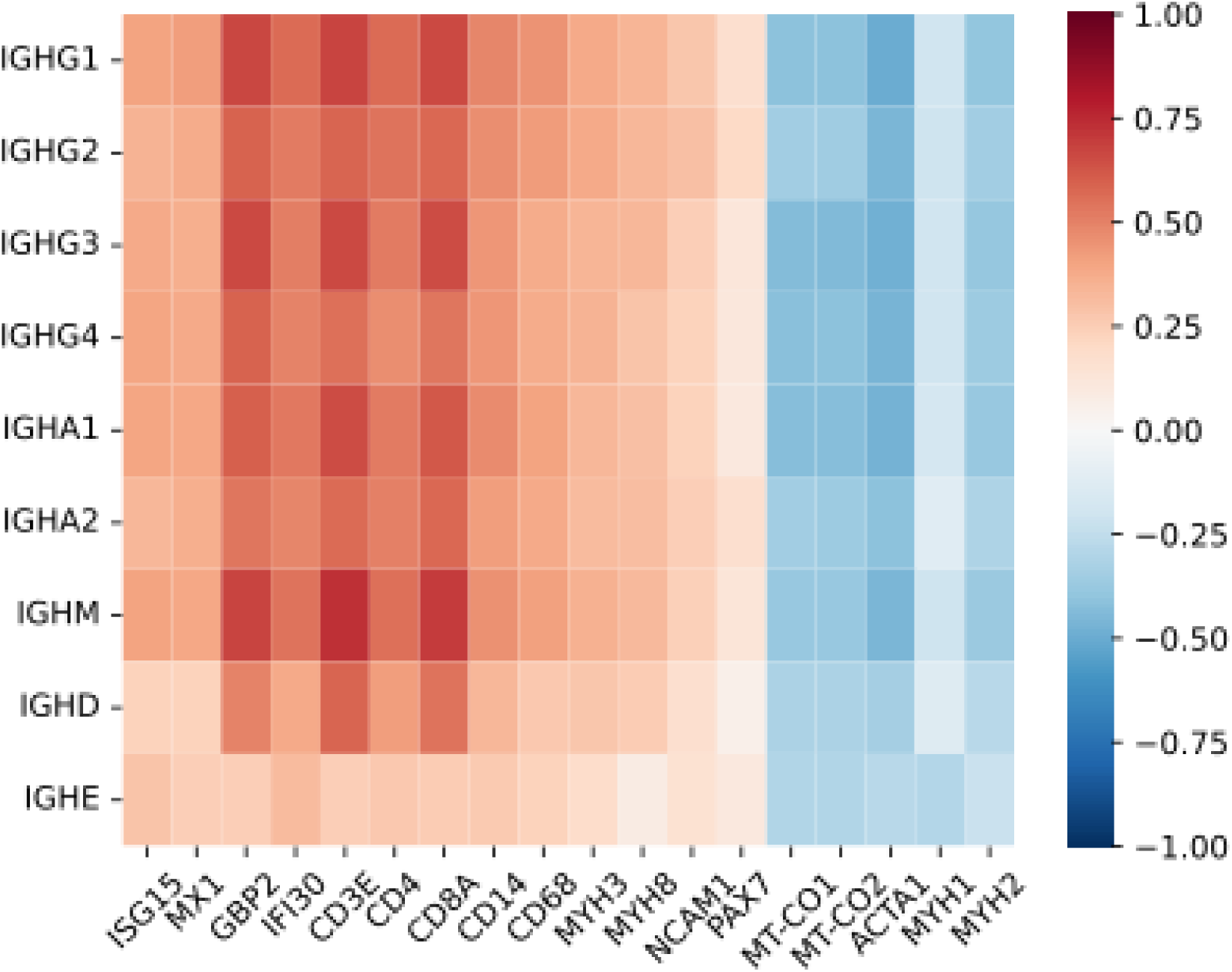
Spearman correlation between immunoglobulin heavy chain gene expression and transcriptomic markers of disease activity across all study samples. Shown are correlations with type I interferon-inducible genes (ISG15, MX1), type II interferon-inducible genes (GBP2, IFI30), T-cell markers (CD3E, CD4, CD8), macrophage markers (CD14, CD68), muscle differentiation markers (MYH3, MYH8, NCAM1, PAX7), mitochondrial genes (MT-CO1, MT-CO2), and structural mature

The strength of these associations varied across clinical and serological subgroups. Anti-Mi2 dermatomyositis showed the strongest correlation between local immunoglobulin expression and disease activity, followed by anti-MDA5, anti-Jo1, and IBM, which demonstrated moderate associations. Anti-TIF1γ and anti-HMGCR subgroups exhibited weaker but consistent correlations, while anti-NXP2 and anti-SRP patients showed minimal association (Supplementary Figure 1). These findings suggest that the contribution of local immunoglobulin expression to disease activity may differ across myositis subtypes.

### Expression of Light Chain and Non-Constant Regions of the Heavy Chain

Analysis of the diversity (D) region of the heavy chain and the joining (J) region of the light chains was not feasible due to insufficient read coverage. In contrast, expression of the constant regions of the light chains, as well as the measurable variable (V) and joining (J) regions of both heavy and light chains, revealed a consistent pattern of preferential overexpression in IBM and ASyS (Supplementary Figures 2–6; Supplementary Tables 2–6). This trend was uniform across all analyzed regions, without evidence of preferential segment usage in specific myositis subgroups.

Expression of different immunoglobulin segments—whether constant or variable—was highly correlated and strongly associated with transcriptomic markers of disease activity (Supplementary Figures 7–8). This included the abovementioned constant regions of different immunoglobulin isotypes and extended to other regions of both heavy and light chains.

Regarding the light chain constant regions, IGKC and IGLC2 were the most abundantly expressed, followed by IGLC3 and IGLC1, while IGLC6 and IGLC7 showed low expression (Supplementary Figure 2; Supplementary Table 2). Among the variable regions, IGHV1-18 was the most highly expressed heavy chain variable segment across all study groups (Supplementary Figure 3; Supplementary Table 3). The heavy chain joining region exhibited low expression overall (Supplementary Figure 4; Supplementary Table 4). For the light chains, IGKV4-1 was the most highly expressed kappa chain variable segment (Supplementary Figure 5; Supplementary Table 5), while several lambda chain variable segments—including IGLV1-40 and IGLV2-23— showed abundant expression (Supplementary Figure 6; Supplementary Table 6).

### Type II Interferon Signaling Drives Local Immunoglobulin Expression

Gene set enrichment analysis of genes correlated with IGHG1 expression revealed a robust association between local immunoglobulin expression and the IFN-γ pathway (Figure 4). Consistent with this, immunoglobulin transcripts correlated tightly with IFN-γ–inducible genes (e.g. GBP2, Figure 5). These results suggest that local immunoglobulin production may be driven by IFN-γ in myositis muscle.

**Figure 4.**
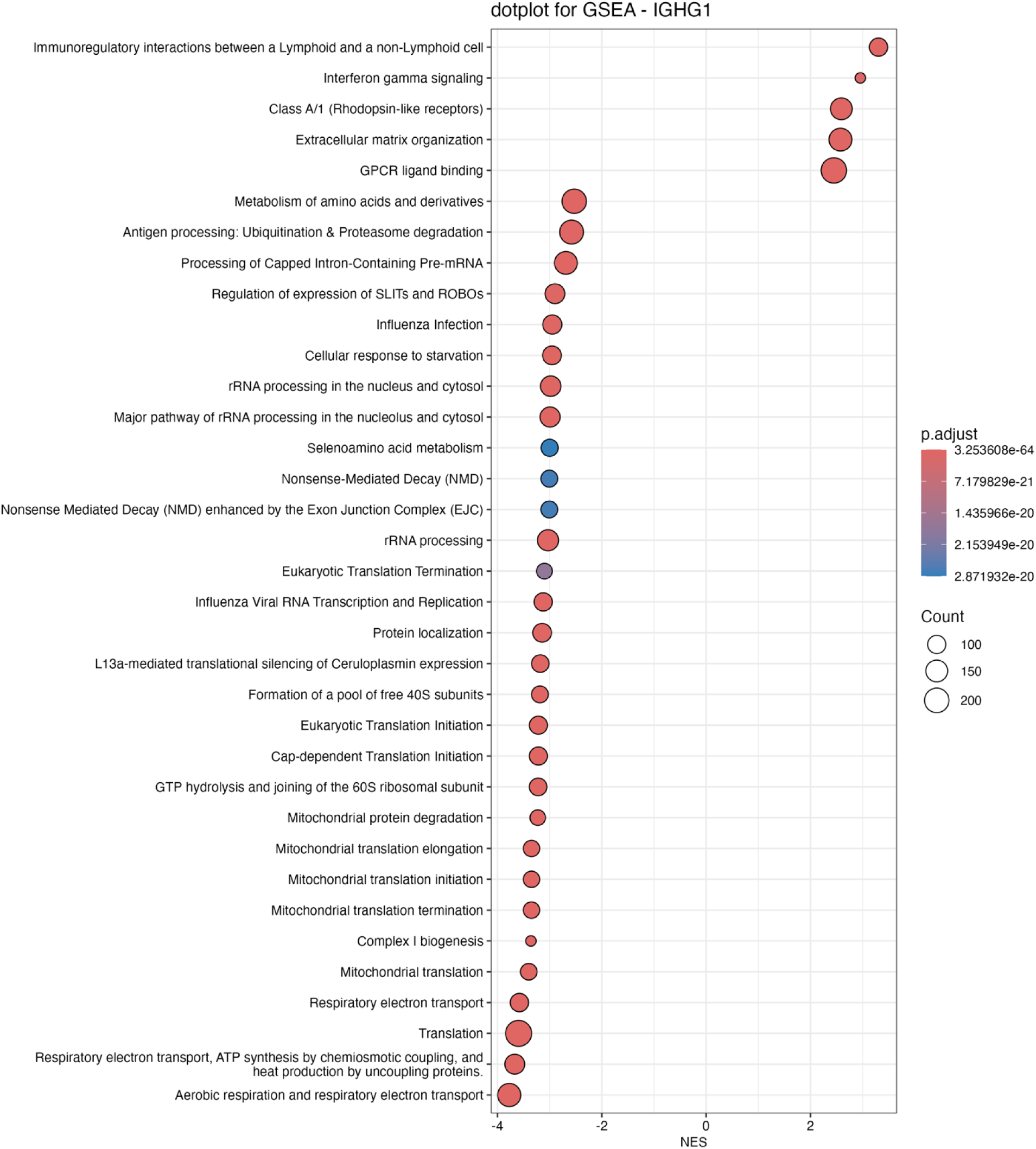
Gene Set Enrichment Analysis (GSEA) of Reactome pathways based on IGHG1 co-expression. This dot plot displays the top 35 Reactome pathways enriched in the Spearman correlation analysis between IGHG1 expression and genome-wide gene expression. The x-axis shows the Normalized Enrichment Score (NES), where positive values indicate pathways positively associated with IGHG1 co-expression and negative values indicate downregulated pathways.

**Figure 5.**
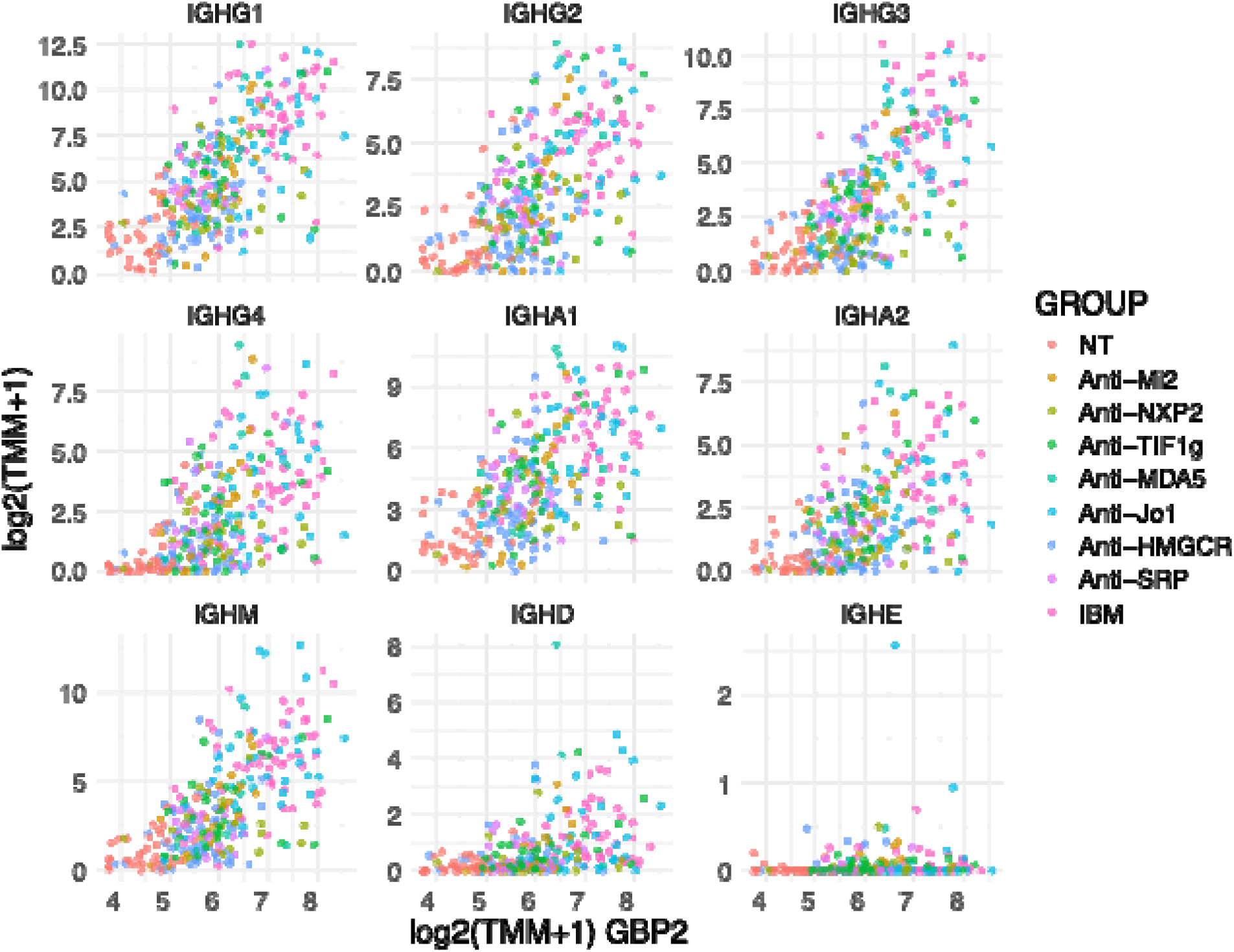
Spearman correlation of the expression of immunoglobulin heavy chain genes with the interferon gamma inducible gene GBP2. Each point represents an individual sample. Expression is presented as trimmed mean of M values (TMM). Normal muscle biopsy: NT; inclusion body myositis: IBM.

### Immunoglobulin Gene Expression Distinguishes Select Myositis Subgroups

We investigated whether immunoglobulin gene expression alone could stratify patients into clinically meaningful subgroups. Dermatomyositis and IMNM did not form distinct clusters. In contrast, samples from patients with ASyS and IBM were clearly separated from healthy muscle biopsies (Figure 6).

**Figure 6.**
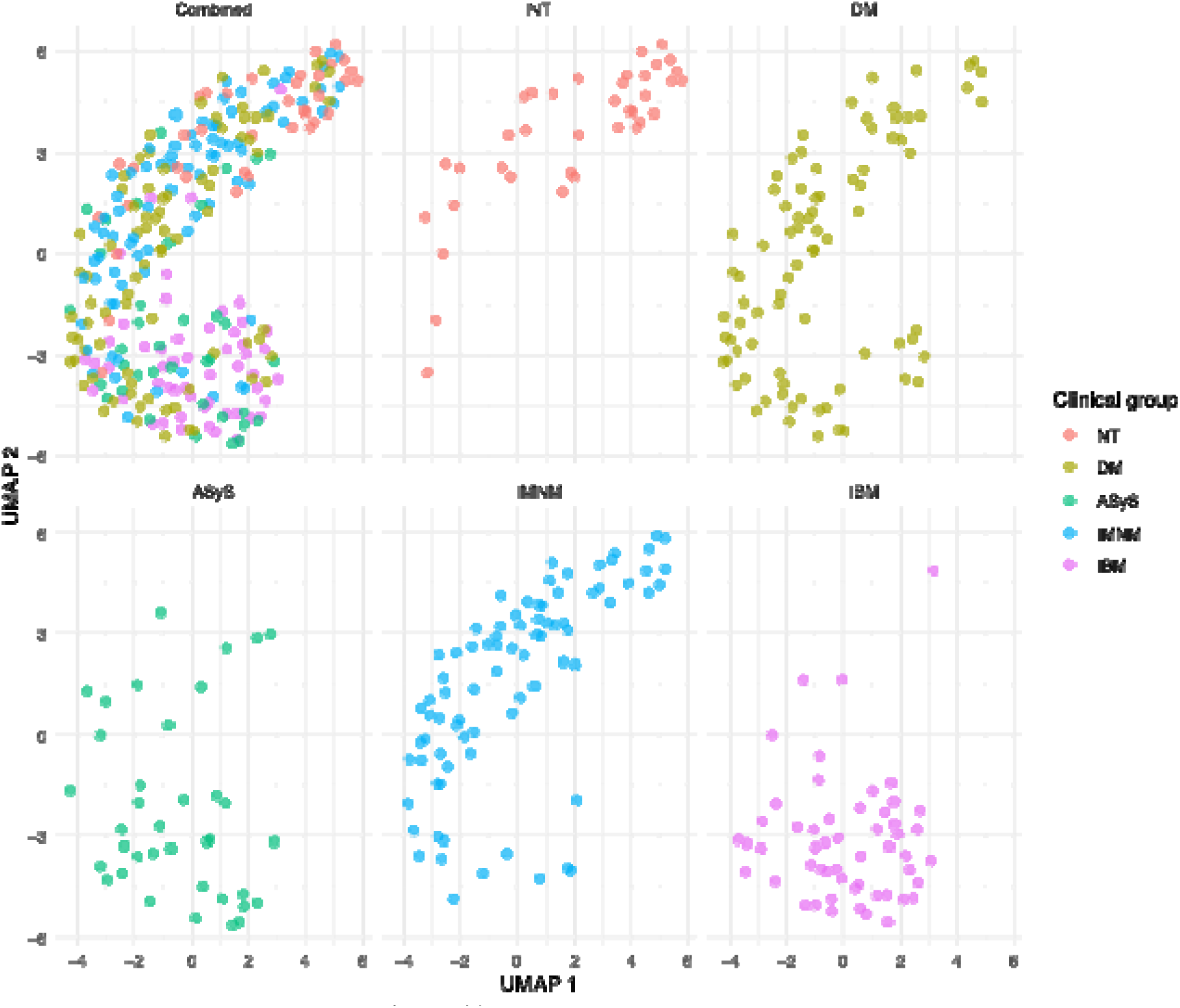
UMAP visualization of clinical groups based on immunoglobulin gene expression. Uniform Manifold Approximation and Projection (UMAP) was used to cluster samples by their immunoglobulin gene expression profiles. Sample groups include normal muscle biopsy (NT), dermatomyositis (DM), antisynthetase syndrome (ASyS), immune-mediated necrotizing myopathy (IMNM), and inclusion body myositis (IBM).

## DISCUSSION

Our data reveal a pronounced intramuscular humoral immune signature in myositis, especially in IBM and ASyS. Consistent with earlier transcriptomic studies in IBM, we found that immunoglobulin gene transcripts are overexpressed genes in affected muscle (3–6). In IBM and ASyS, transcripts of heavy chains for IgG, IgA and IgM were markedly elevated, reflecting infiltration by class-switched B cells or plasma cells. This local immunoglobulin expression closely tracked clinical severity, suggesting it could serve as a biomarker of disease activity. Indeed, unsupervised clustering by Ig-gene expression cleanly separated IBM and ASyS muscles from controls, underscoring its potential utility in stratifying myositis subtypes.

A striking feature was the strong association of the local Ig transcript signature with a type II interferon (IFN-γ) gene program. Recent analyses have shown that IBM and ASyS muscle are characterized by a predominance of IFN-γ–induced transcripts (12, 15). At the cellular level, IFN-γ is primarily produced by infiltrating cytotoxic T cells in IBM/ASyS, which in turn acts on muscle fibers and resident immune cells. Of note, IFN-γ is a key factor to induce the migration of B cells from the lymph nodes to the tissues as tissue-resident B cells (17) that will survive in the tissues if they receive survival signals by continuous antigenic stimulation. This could explain why, in myositis, B-cells are preferentially attracted to tissues where there is expression of their cognate autoantigens. Also, IFN-γ signaling directly promotes plasma cell differentiation in muscle and other tissues (18, 19). Such a mechanism would explain the high levels of intramuscular Ig transcription seen in IBM and ASyS.

The identification of a robust IFN-γ–associated Ig signature in IBM and ASyS has several clinical ramifications. First, it offers a biomarker for disease stratification and activity assessment. Muscle biopsies showing elevated immunoglobulin transcripts might help distinguish IBM/ASyS from other inflammatory myopathies. Serial measurement of this signature could also monitor response to therapy: for example, declining local Ig gene expression might indicate therapeutic suppression of B-cell activity.

Second, our findings reinforce the rationale for therapies targeting autoantibodies and B cells in myositis. Intravenous immunoglobulin, which saturates the neonatal Fc receptor (FcRn) and reduces immunoglobulin half-life, has consistently shown efficacy across multiple forms of myositis (20, 21). This same mechanism is being leveraged by synthetic FcRn inhibitors, such as efgartigimod, which have demonstrated benefit in other autoantibody-mediated diseases (22) and may similarly prove effective in myositis (23, 24).

In parallel, therapies that deplete B cells (e.g., rituximab) or target B cells and plasma cells more broadly have shown benefit in various myositis subtypes. Although the largest clinical trial of rituximab in myositis did not meet its primary endpoint (25), subsequent case series (26), post-hoc analysis (27), and extensive clinical experience support its effectiveness in reducing autoantibody titers and improving outcomes in refractory patients. Our findings may help account for the limited efficacy of rituximab reported in some studies given that the drug may not effectively target tissue-resident B cells as it does circulating ones.

A more recent and promising strategy is autologous CD19-directed CAR-T cell therapy, which has induced rapid and sustained remissions in patients with refractory disease. For instance, in three ASyS patients, CAR-T cell therapy led to rapid clinical improvement, seroconversion to autoantibody negativity, normalization of creatine kinase levels, and durable recovery of muscle strength (28). Notably, the only patient who did not seroconvert relapsed after 9 months but achieved drug-free remission following anti-BCMA therapy (29). These striking results underscore the central pathogenic role of B cells in ASyS and support the idea that eliminating autoantibody-producing cells may reset the autoimmune response. Our data further support this therapeutic direction by highlighting substantial local B-cell activity in the muscle tissue of myositis patients.

The present study has limitations. Because we used bulk RNA sequencing on heterogeneous muscle biopsies, we cannot definitively assign Ig transcripts to specific cell types (e.g. resident plasma cells vs. infiltrating B cells). Nor can we reconstruct full B-cell receptor sequences, as our data lack complete V(D)J coverage. Also, immunoglobulin gene segments that differ substantially from the reference genome may not have been fully captured or analyzed; however, any resulting bias would likely affect all groups equally. Although this study does not provide functional validation of the pathogenic role of locally expressed autoantibodies, such evidence has been demonstrated previously for several myositis-associated autoantibodies (2). Likewise, the results are restricted to the transcriptomic measurement and did not explore the abundance at the protein level.

Future studies should address these gaps. Single-cell RNA sequencing (with VDJ repertoire analysis) of muscle-infiltrating immune cells could pinpoint the exact B-cell subsets producing the immunoglobulin transcripts and reveal clonal expansions. Spatial transcriptomics or multiplex immunofluorescence could map the cellular interactions among IFN-γ–producing T cells, and plasma cells. *In vitro* co-culture of human myotubes with immune cells under IFN-γ stimulation could test causality of B-cell recruitment and differentiation. Most importantly, future interventional studies should investigate whether inhibiting IFN-γ signaling can reduce intramuscular antibody production and improve clinical outcomes, particularly in myositis subtypes lacking effective treatments, such as IBM.

In summary, our findings demonstrate that myositis—particularly IBM and ASyS—is characterized by prominent local immunoglobulin gene expression that is driven by an IFN-γ–rich inflammatory environment and associated with disease activity. This links cellular and humoral arms of the immune response in myositis and provides a molecular basis for tailored biomarker and therapeutic strategies.

## Data Availability

All data produced in the present study are available upon reasonable request to the authors

## Acknowledgments

Special thanks to Julie Thompson for her invaluable help maintaining the NIH Natural History Protocol, the NIAMS Sequencing Core, and its members.

---

**Supplementary Figure 1.**
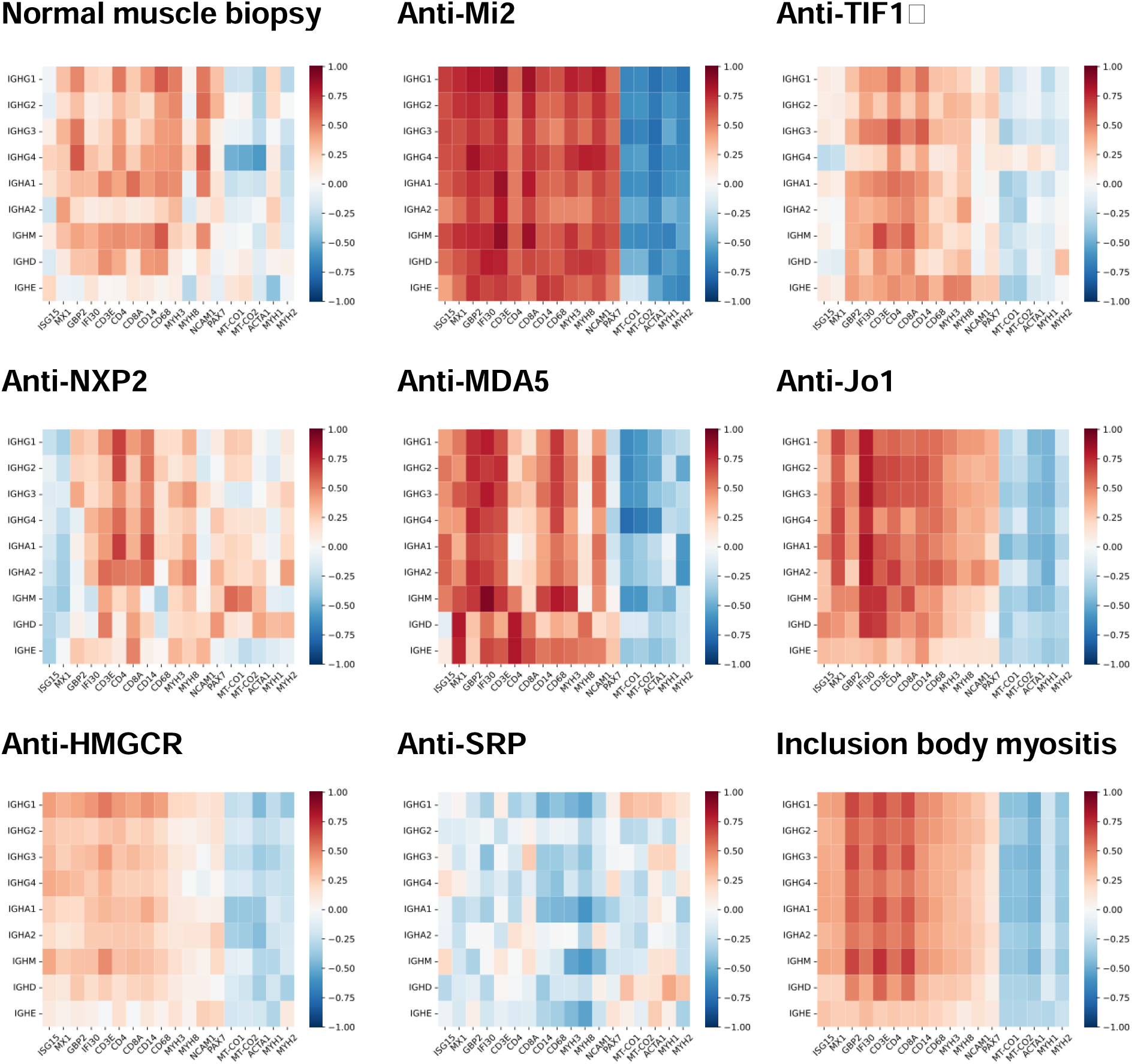
Spearman correlations between immunoglobulin heavy chain gene expression and markers of disease activity across study groups. Shown are correlations with type I interferon-inducible genes (ISG15, MX1), type II interferon-inducible genes (GBP2, IFI30), T-cell markers (CD3E, CD4, CD8), macrophage markers (CD14, CD68), muscle differentiation markers (MYH3, MYH8, NCAM1, PAX7), mitochondrial genes (MT-CO1, MT-CO2), and structural mature muscle proteins (ACTA1, MYH1, MYH2).

**Supplementary Figure 2.**
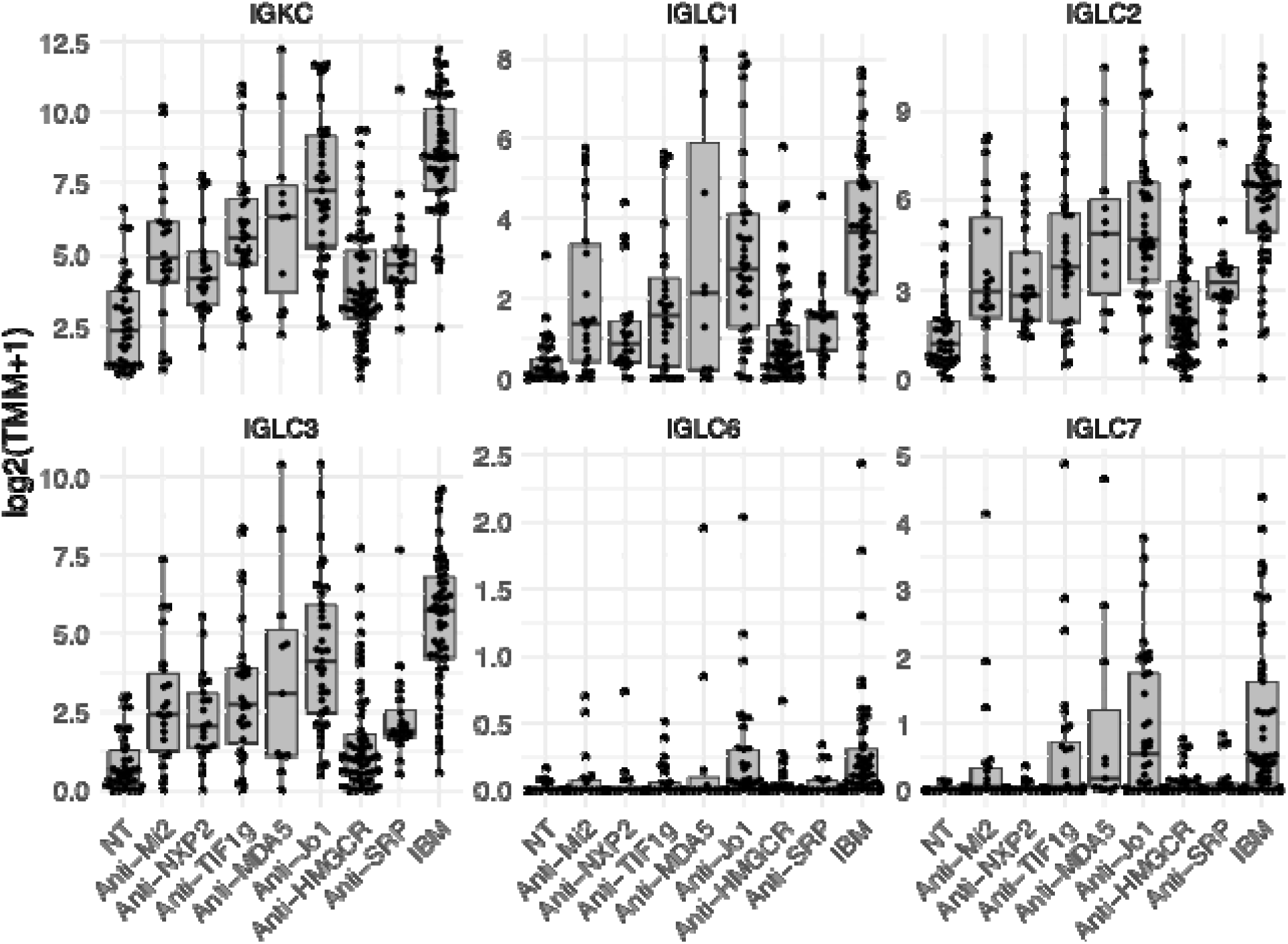
Expression of immunoglobulin kappa and lambda light chain constant region genes across disease groups. Each point represents an individual sample. Expression is presented as trimmed mean of M values (TMM). Normal muscle biopsy: NT; inclusion body myositis: IBM.

**Supplementary Figure 3.**
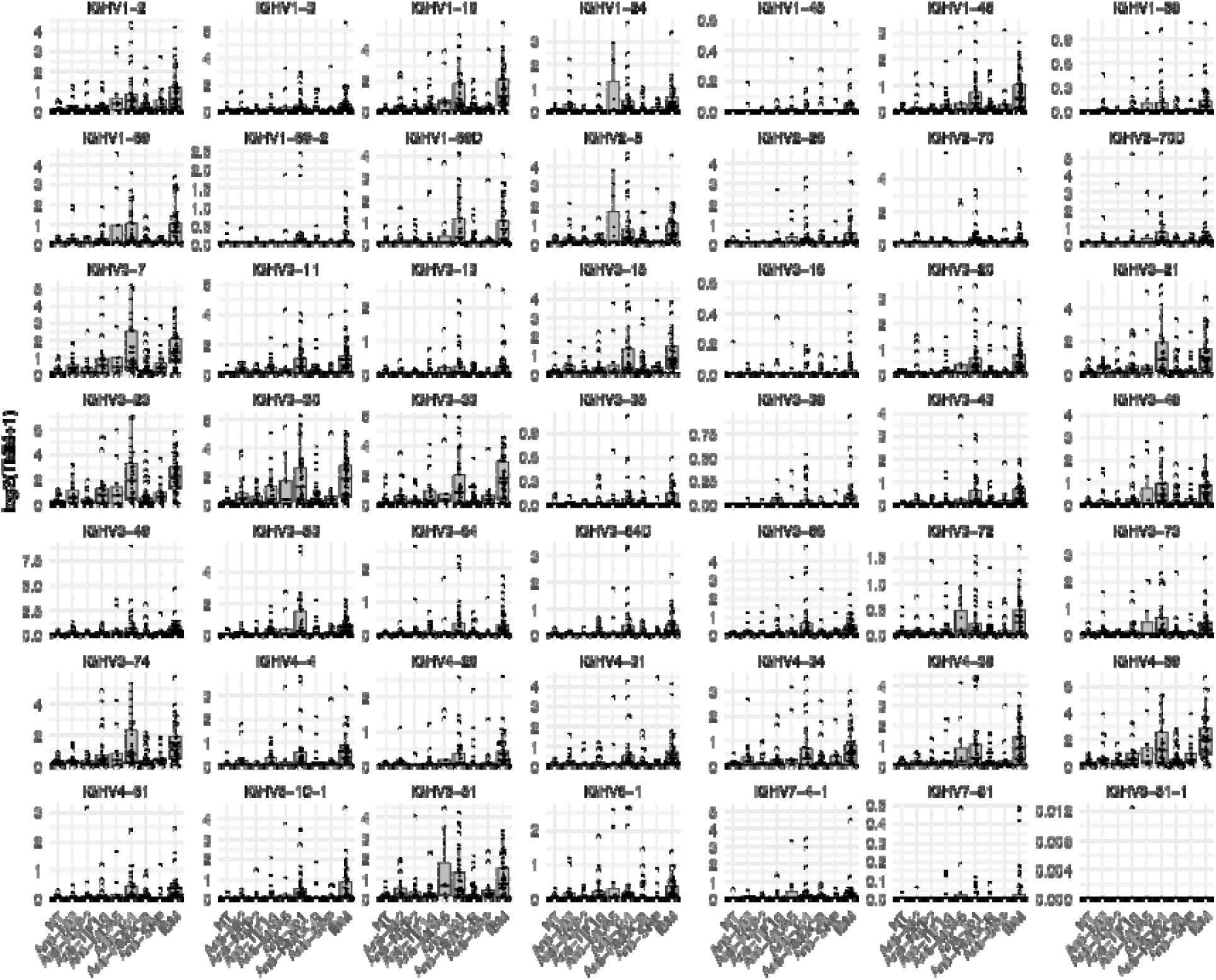
Expression of immunoglobulin heavy chain variable region genes across disease groups. Each point represents an individual sample. Expression is presented as trimmed mean of M values (TMM). Normal muscle biopsy: NT; inclusion body myositis: IBM.

**Supplementary Figure 4.**
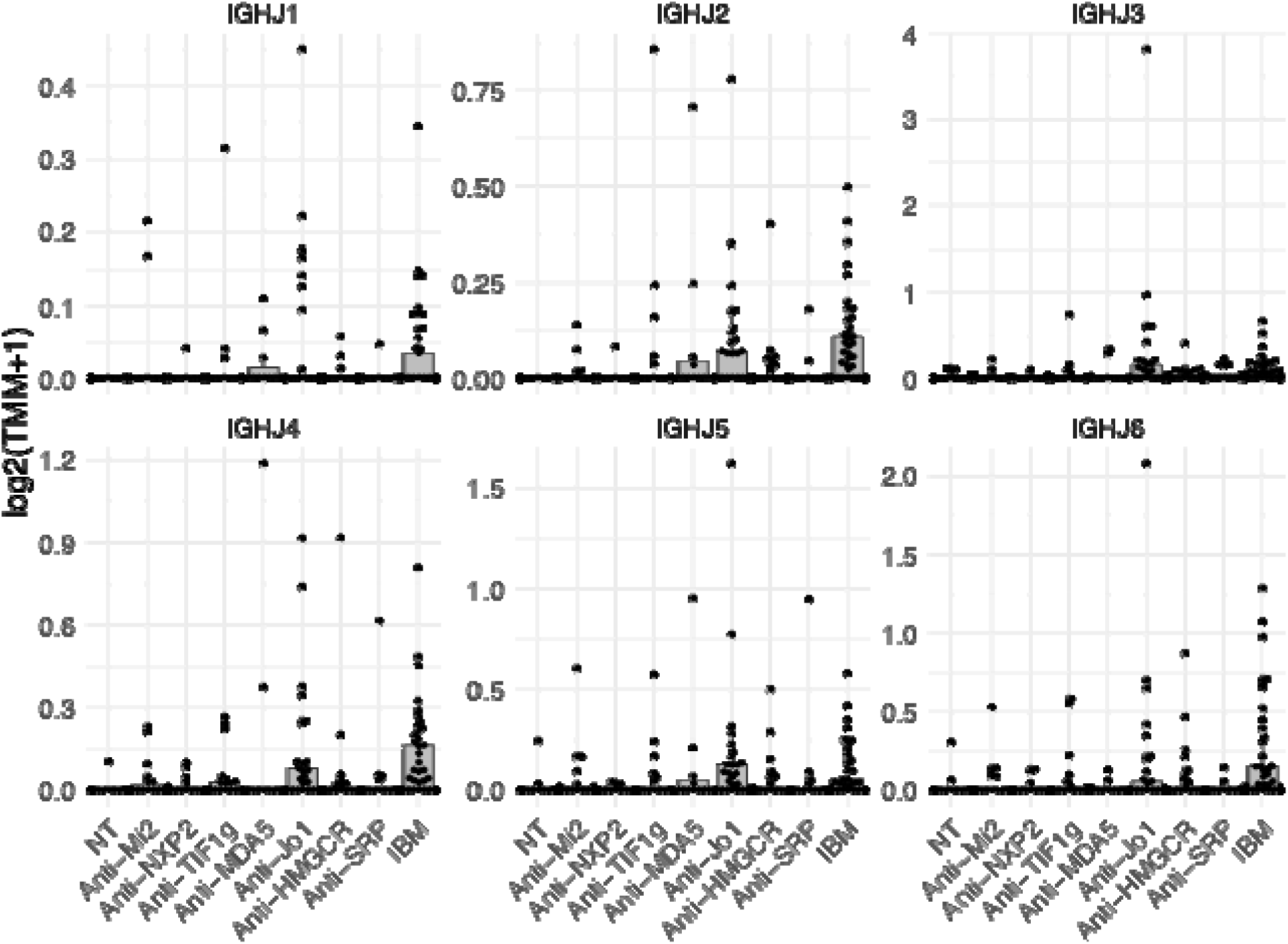
Expression of immunoglobulin heavy chain joining region genes across disease groups. Each point represents an individual sample. Expression is presented as trimmed mean of M values (TMM). Normal muscle biopsy: NT; inclusion body myositis: IBM.

**Supplementary Figure 5.**
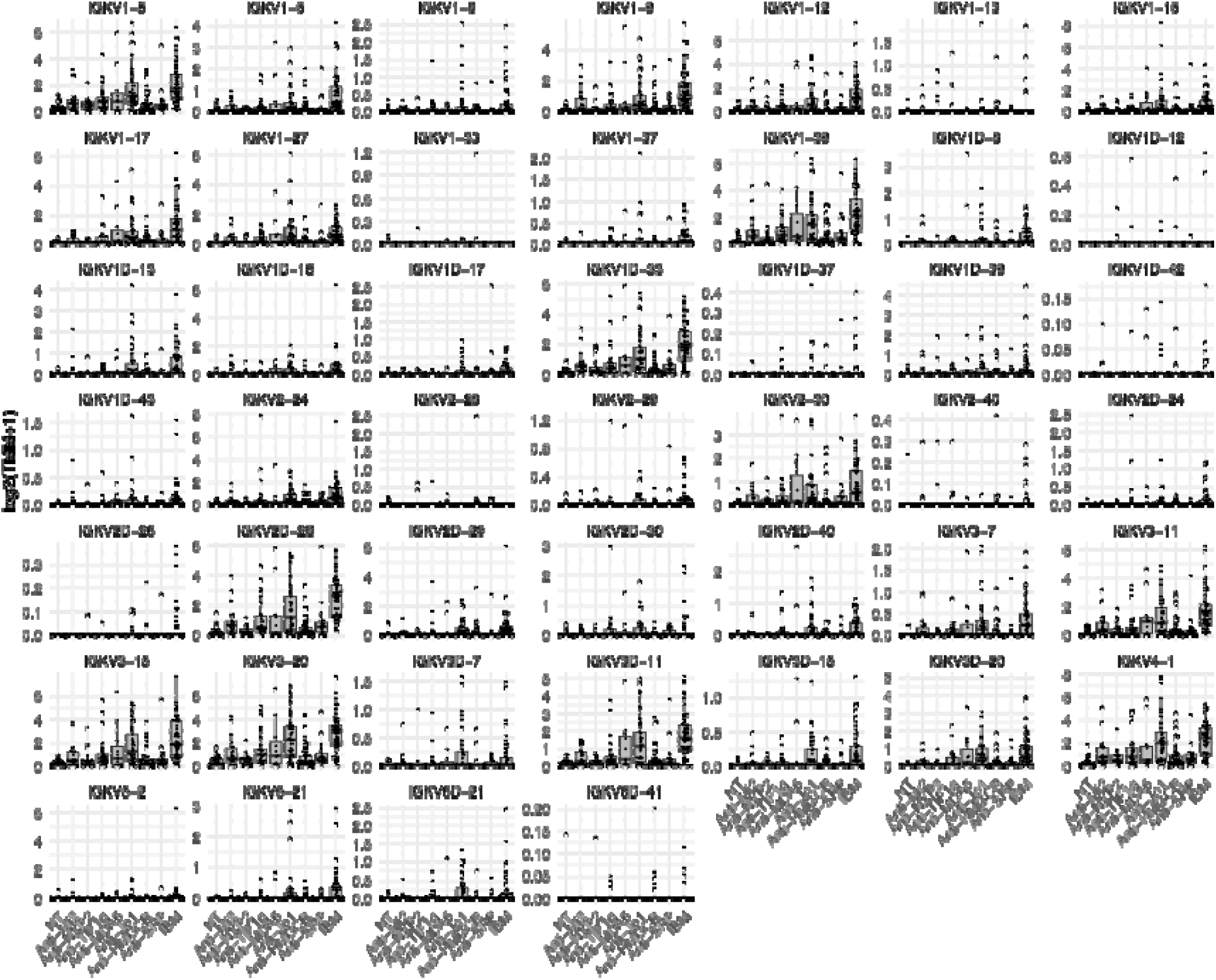
Expression of immunoglobulin kappa light chain variable region genes across disease groups. Each point represents an individual sample. Expression is presented as trimmed mean of M values (TMM). Normal muscle biopsy: NT; inclusion body myositis: IBM.

**Supplementary Figure 6.**
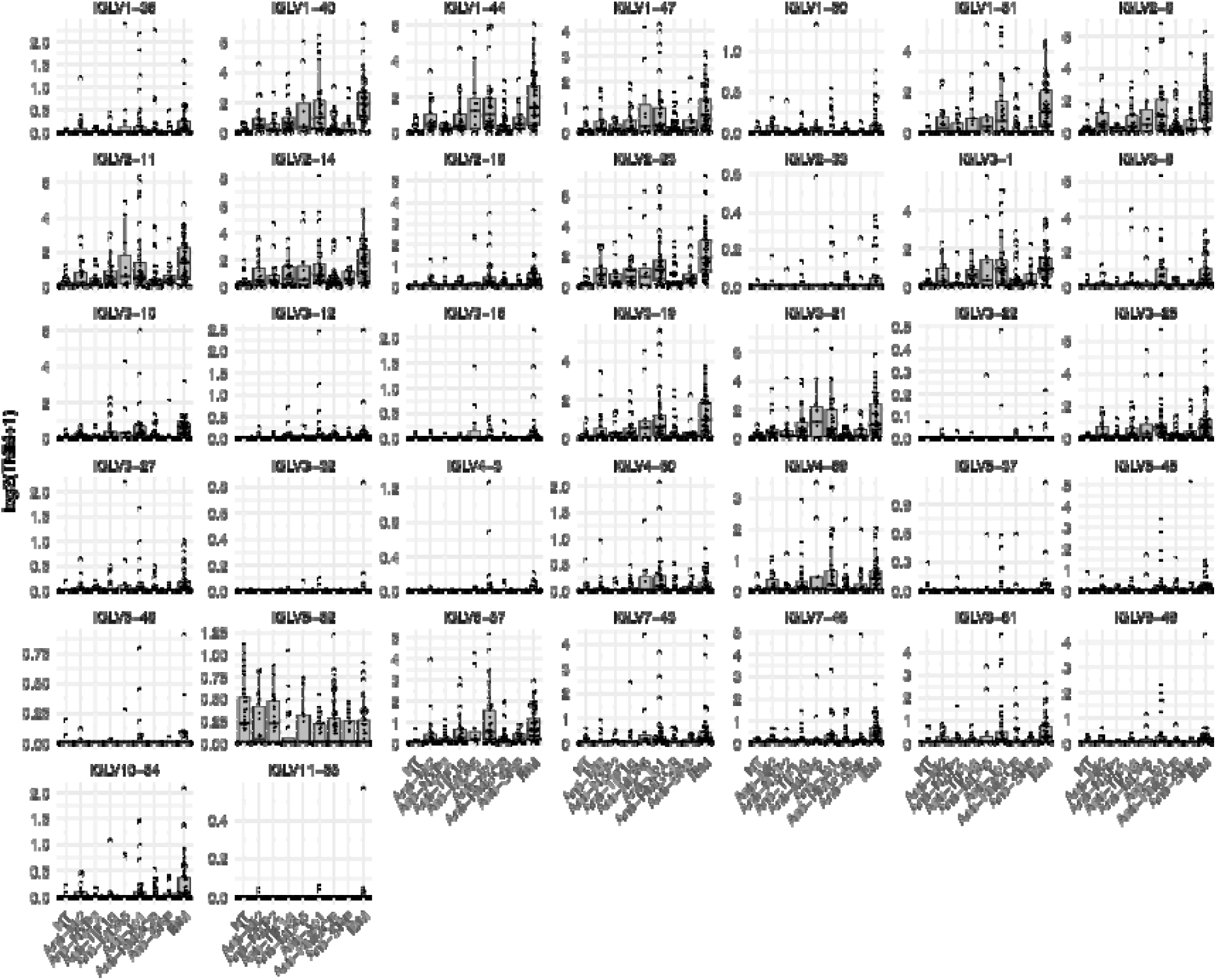
Expression of immunoglobulin lambda light chain variable region genes across disease groups. Each point represents an individual sample. Expression is presented as trimmed mean of M values (TMM). Normal muscle biopsy: NT; inclusion body myositis: IBM.

**Supplementary Figure 7.**
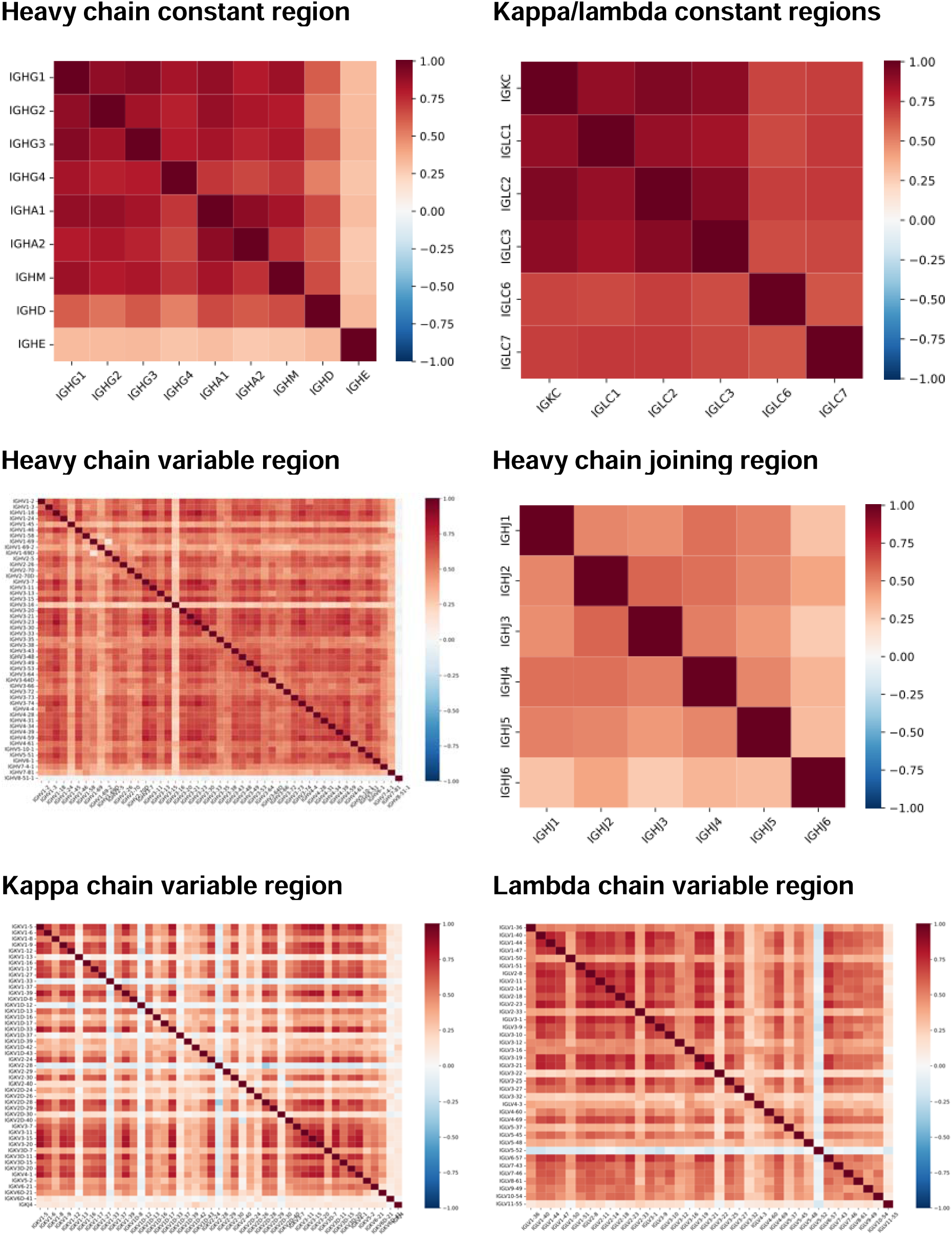
Spearman correlation matrix of immunoglobulin genes.

**Supplementary Figure 8.**
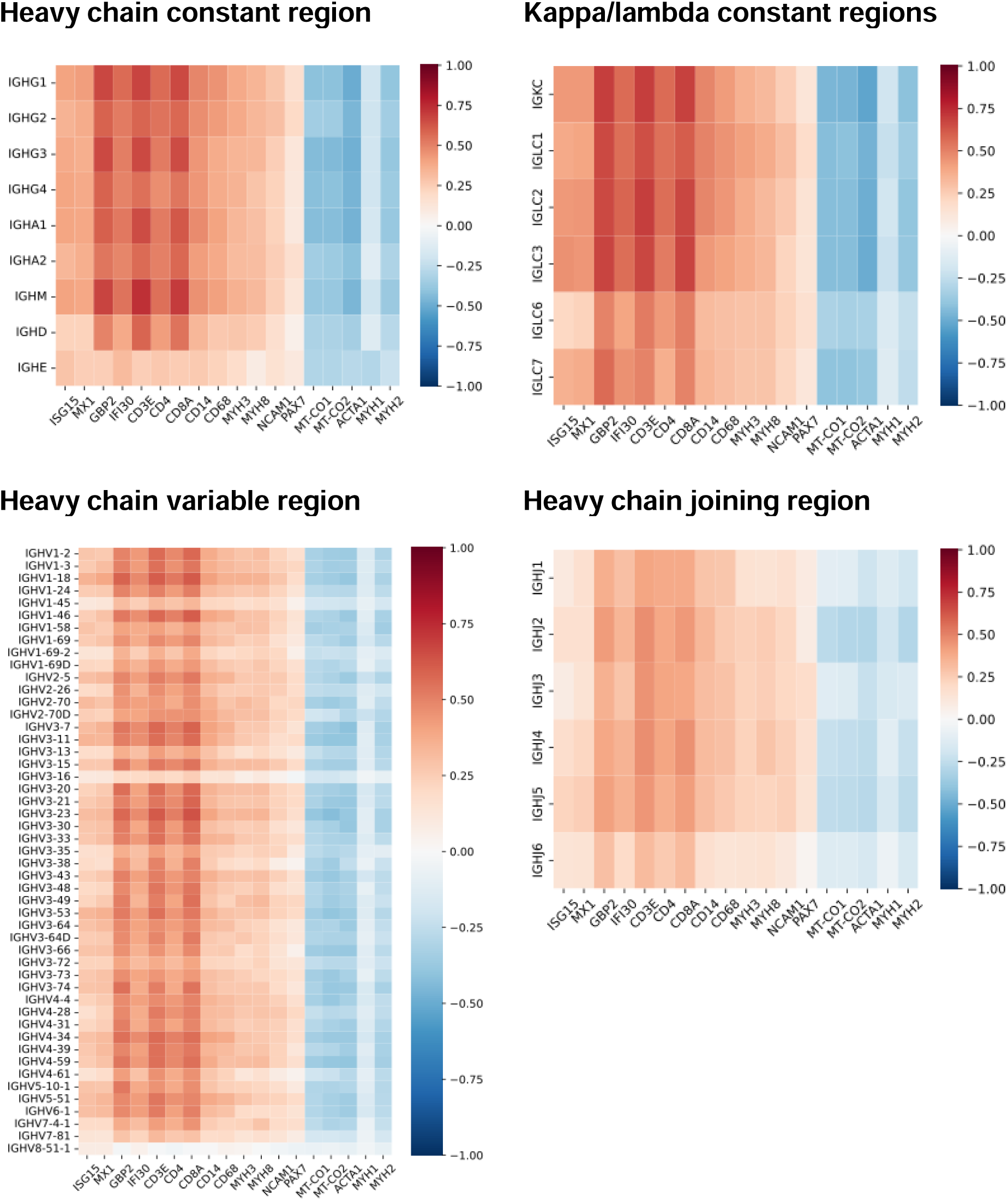
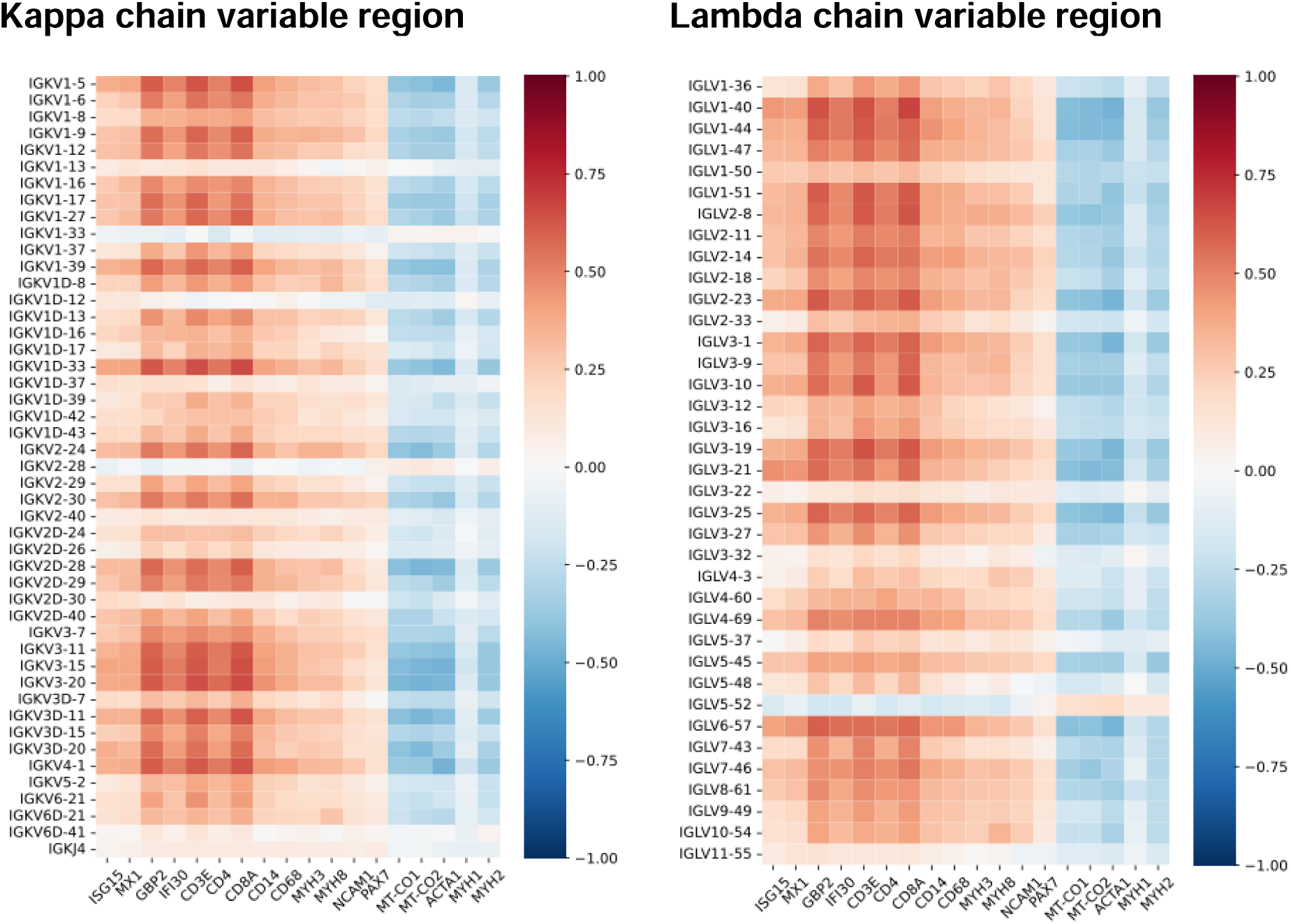
Spearman correlation between immunoglobulin gene expression and markers of disease activity. Shown are correlations with type I interferon-inducible genes (ISG15, MX1), type II interferon-inducible genes (GBP2, IFI30), T-cell markers (CD3E, CD4, CD8), macrophage markers (CD14, CD68), muscle differentiation markers (MYH3, MYH8, NCAM1, PAX7), mitochondrial genes (MT-CO1, MT-CO2), and structural mature muscle proteins (ACTA1, MYH1, MYH2).

**Supplementary Table 1.** Distribution of groups included in the study.

| **Group** | **N = 289<sup>1</sup>** |
| --- | --- |
| NT | 37 (13%) |
| Anti-Mi2 | 22 (7.6%) |
| Anti-NXP2 | 21 (7.3%) |
| Anti-TIF1γ | 28 (9.7%) |
| Anti-MDA5 | 11 (3.8%) |
| Anti-Jo1 | 37 (13%) |
| Anti-HMGCR | 60 (21%) |
| Anti-SRP | 20 (6.9%) |
| IBM | 53 (18%) |

<sup>1</sup>n (%)

**Supplementary Table 2.** Expression of immunoglobulin kappa/lambda constant region genes across disease groups, shown as comparisons versus all other muscle biopsies (vs. ALL) and versus normal muscle biopsies (vs. NT). DM: dermatomyositis; IMNM: immune-mediated necrotizing myopathy; IBM: inclusion body myositis; NT: normal muscle biopsy.

| DM vs. ALL | | | Mi2 vs. ALL | | | NXP2 vs. ALL | | | MDA5 vs. ALL | | | TIF1γvs. ALL | | |
| --- | --- | --- | --- | --- | --- | --- | --- | --- | --- | --- | --- | --- | --- | --- |
| Gene | LogFC | q-val | Gene | LogFC | q-val | Gene | LogFC | q-val | Gene | LogFC | q-val | Gene | LogFC | q-val |
| IGLC6 | -0.7 | 0.024 | IGLC6 | -0.7 | 0.265 | IGLC7 | -1.3 | 0.208 | IGLC2 | 1.5 | 0.587 | IGLC6 | -0.6 | 0.372 |
| IGKC | -0.2 | 0.693 | IGKC | -0.4 | 0.691 | IGLC6 | -0.8 | 0.291 | IGLC7 | 1.2 | 0.701 | IGLC7 | 0.4 | 0.714 |
| IGLC7 | -0.2 | 0.767 | IGLC1 | 0.4 | 0.767 | IGKC | -1.0 | 0.481 | IGLC3 | 1.0 | 0.722 | IGKC | 0.3 | 0.762 |
| IGLC2 | 0.1 | 0.832 | IGLC2 | -0.3 | 0.775 | IGLC1 | -0.8 | 0.658 | IGKC | 0.8 | 0.739 | IGLC2 | 0.3 | 0.797 |
| IGLC3 | 0.1 | 0.840 | IGLC7 | -0.3 | 0.804 | IGLC3 | -0.5 | 0.740 | IGLC1 | 0.7 | 0.812 | IGLC3 | 0.3 | 0.835 |
| IGLC1 | -0.1 | 0.899 | IGLC3 | 0.0 | 0.995 | IGLC2 | -0.4 | 0.762 | IGLC6 | 0.2 | 0.939 | IGLC1 | -0.2 | 0.892 |

| JO1 vs. ALL |  |  | IMNM vs. ALL |  |  | HMGCR vs. ALL |  |  | SRP vs. ALL |  |  | IBM vs. ALL |  |  |
| --- | --- | --- | --- | --- | --- | --- | --- | --- | --- | --- | --- | --- | --- | --- |
| Gene | LogFC | q-val | Gene | LogFC | q-val | Gene | LogFC | q-val | Gene | LogFC | q-val | Gene | LogFC | q-val |
| IGKC | 2.1 | 3.45e-04 | IGLC7 | -1.8 | 1.46e-04 | IGLC7 | -1.8 | 5.74e-04 | IGLC7 | -0.9 | 0.996 | IGKC | 3.8 | 1.26e-24 |
| IGLC3 | 2.2 | 8.79e-04 | IGKC | -1.9 | 0.001 | IGLC3 | -2.4 | 0.003 | IGKC | -0.7 | 0.996 | IGLC3 | 3.8 | 1.10e-20 |
| IGLC7 | 2.1 | 0.001 | IGLC3 | -2.2 | 0.001 | IGKC | -2.0 | 0.004 | IGLC6 | -0.3 | 0.996 | IGLC1 | 3.7 | 5.40e-16 |
| IGLC2 | 2.0 | 0.003 | IGLC2 | -1.8 | 0.003 | IGLC2 | -2.0 | 0.005 | IGLC3 | -0.4 | 0.996 | IGLC2 | 3.1 | 7.35e-15 |
| IGLC1 | 2.2 | 0.004 | IGLC6 | -0.8 | 0.012 | IGLC6 | -0.9 | 0.012 | IGLC2 | -0.3 | 0.996 | IGLC7 | 2.6 | 6.01e-09 |
| IGLC6 | 1.0 | 0.130 | IGLC1 | -1.8 | 0.017 | IGLC1 | -2.2 | 0.017 | IGLC1 | 0.0 | 0.999 | IGLC6 | 1.7 | 3.94e-07 |

| DM vs. NT | | | Mi2 vs. NT | | | NXP2 vs. NT | | | MDA5 vs. NT | | | TIF1γvs. NT | | |
| --- | --- | --- | --- | --- | --- | --- | --- | --- | --- | --- | --- | --- | --- | --- |
| Gene | LogFC | q-val | Gene | LogFC | q-val | Gene | LogFC | q-val | Gene | LogFC | q-val | Gene | LogFC | q-val |
| IGKC | 3.1 | 6.28e-05 | IGLC3 | 3.6 | 7.39e-05 | IGLC3 | 3.1 | 1.79e-05 | IGLC2 | 4.5 | 2.31e-05 | IGKC | 3.5 | 9.68e-06 |
| IGLC3 | 3.7 | 7.30e-05 | IGKC | 2.8 | 2.46e-04 | IGLC2 | 2.7 | 9.99e-05 | IGKC | 3.9 | 1.64e-04 | IGLC2 | 3.3 | 2.84e-04 |
| IGLC2 | 3.2 | 2.25e-04 | IGLC1 | 3.9 | 2.49e-04 | IGKC | 2.3 | 1.75e-04 | IGLC3 | 4.5 | 8.89e-04 | IGLC7 | 2.2 | 5.43e-04 |
| IGLC7 | 1.7 | 3.31e-04 | IGLC2 | 2.8 | 0.003 | IGLC1 | 2.8 | 0.001 | IGLC7 | 2.9 | 0.001 | IGLC3 | 3.8 | 6.17e-04 |
| IGLC1 | 3.5 | 6.19e-04 | IGLC7 | 1.5 | 0.007 | IGLC7 | 0.6 | 0.154 | IGLC1 | 4.2 | 0.004 | IGLC1 | 3.4 | 0.003 |
| IGLC6 | 0.2 | 0.671 | IGLC6 | -0.1 | 0.906 | IGLC6 | -0.1 | 0.900 | IGLC6 | 0.7 | 0.321 | IGLC6 | 0.1 | 0.737 |

| JO1 vs. NT |  |  | IMNM vs. NT |  |  | HMGCR vs. NT |  |  | SRP vs. NT |  |  | IBM vs. NT |  |  |
| --- | --- | --- | --- | --- | --- | --- | --- | --- | --- | --- | --- | --- | --- | --- |
| Gene | LogFC | q-val | Gene | LogFC | q-val | Gene | LogFC | q-val | Gene | LogFC | q-val | Gene | LogFC | q-val |
| IGLC7 | 3.7 | 2.53e-09 | IGKC | 1.8 | 0.002 | IGKC | 1.6 | 0.010 | IGLC1 | 3.5 | 2.12e-06 | IGLC7 | 4.0 | 1.92e-11 |
| IGKC | 5.1 | 2.49e-08 | IGLC1 | 2.2 | 0.009 | IGLC1 | 1.8 | 0.039 | IGLC2 | 2.8 | 6.68e-06 | IGLC1 | 6.6 | 1.17e-09 |
| IGLC3 | 5.5 | 3.81e-07 | IGLC2 | 1.8 | 0.010 | IGLC2 | 1.5 | 0.044 | IGLC3 | 3.2 | 1.07e-05 | IGKC | 6.4 | 1.45e-09 |
| IGLC2 | 4.8 | 5.07e-07 | IGLC3 | 2.0 | 0.011 | IGLC3 | 1.7 | 0.045 | IGKC | 2.6 | 2.23e-05 | IGLC6 | 2.1 | 1.77e-08 |
| IGLC1 | 5.5 | 2.68e-05 | IGLC7 | 0.6 | 0.109 | IGLC7 | 0.4 | 0.261 | IGLC7 | 1.0 | 0.051 | IGLC3 | 6.8 | 1.92e-08 |
| IGLC6 | 1.4 | 2.54e-04 | IGLC6 | 0.0 | 0.910 | IGLC6 | -0.1 | 0.813 | IGLC6 | 0.5 | 0.196 | IGLC2 | 5.8 | 5.41e-07 |

**Supplementary Table 3.** Expression of top 10 immunoglobulin heavy chain variable region genes across disease groups, shown as comparisons versus all other muscle biopsies (vs. ALL) and versus normal muscle biopsies (vs. NT). DM: dermatomyositis; IMNM: immune-mediated necrotizing myopathy; IBM: inclusion body myositis; NT: normal muscle biopsy.

| DM vs. ALL |  |  | Mi2 vs. ALL |  |  | NXP2 vs. ALL |  |  | MDA5 vs. ALL |  |  | TIF1γvs. ALL |  |  |
| --- | --- | --- | --- | --- | --- | --- | --- | --- | --- | --- | --- | --- | --- | --- |
| Gene | LogFC | q-val | Gene | LogFC | q-val | Gene | LogFC | q-val | Gene | LogFC | q-val | Gene | LogFC | q-val |
| IGHV1-69-2 | -0.8 | 0.003 | IGHV3-38 | -1.1 | 0.009 | IGHV4-28 | -1.3 | 0.030 | IGHV5-51 | 1.8 | 0.448 | IGHV1-69-2 | -0.9 | 0.087 |
| IGHV3-38 | -0.6 | 0.006 | IGHV7-81 | -0.8 | 0.018 | IGHV1-24 | -1.4 | 0.034 | IGHV1-2 | 1.3 | 0.649 | IGHV1-45 | -0.5 | 0.177 |
| IGHV3-16 | -0.5 | 0.007 | IGHV4-61 | -1.2 | 0.032 | IGHV4-4 | -1.5 | 0.034 | IGHV1-24 | 1.1 | 0.694 | IGHV3-16 | -0.5 | 0.250 |
| IGHV1-45 | -0.4 | 0.012 | IGHV1-69-2 | -1.0 | 0.035 | IGHV3-20 | -1.5 | 0.035 | IGHV1-69 | 1.2 | 0.700 | IGHV1-24 | -0.8 | 0.259 |
| IGHV2-26 | -0.8 | 0.013 | IGHV3-35 | -0.8 | 0.039 | IGHV2-26 | -1.2 | 0.072 | IGHV2-5 | 1.1 | 0.701 | IGHV7-4-1 | -0.7 | 0.309 |
| IGHV7-81 | -0.4 | 0.018 | IGHV2-26 | -1.2 | 0.057 | IGHV1-69 | -1.5 | 0.084 | IGHV6-1 | 0.9 | 0.716 | IGHV1-69D | -0.9 | 0.313 |
| IGHV3-48 | -0.9 | 0.025 | IGHV1-45 | -0.6 | 0.079 | IGHV3-48 | -1.3 | 0.129 | IGHV3-33 | 1.1 | 0.717 | IGHV4-28 | -0.6 | 0.372 |
| IGHV3-35 | -0.5 | 0.027 | IGHV3-16 | -0.6 | 0.081 | IGHV1-58 | -0.8 | 0.130 | IGHV4-34 | 0.9 | 0.719 | IGHV3-13 | -0.5 | 0.383 |
| IGHV3-64D | -0.7 | 0.032 | IGHV3-64D | -0.9 | 0.111 | IGHV3-64D | -1.0 | 0.153 | IGHV3-13 | 0.8 | 0.719 | IGHV2-5 | -0.7 | 0.422 |
| IGHV3-43 | -0.8 | 0.032 | IGHV3-13 | -0.8 | 0.123 | IGHV4-59 | -1.7 | 0.168 | IGHV3-49 | 0.9 | 0.721 | IGHV4-61 | -0.5 | 0.516 |

| JO1 vs. ALL |  |  | IMNM vs. ALL |  |  | HMGCR vs. ALL |  |  | SRP vs. ALL |  |  | IBM vs. ALL |  |  |
| --- | --- | --- | --- | --- | --- | --- | --- | --- | --- | --- | --- | --- | --- | --- |
| Gene | LogFC | q-val | Gene | LogFC | q-val | Gene | LogFC | q-val | Gene | LogFC | q-val | Gene | LogFC | q-val |
| IGHV3-23 | 2.8 | 9.65e-07 | IGHV3-73 | -1.2 | 3.31e-04 | IGHV3-53 | -1.5 | 5.68e-04 | IGHV3-35 | -0.6 | 0.928 | IGHV4-59 | 3.6 | 5.44e-19 |
| IGHV3-7 | 2.6 | 1.69e-06 | IGHV3-53 | -1.4 | 4.95e-04 | IGHV3-74 | -1.9 | 0.001 | IGHV3-64 | -0.8 | 0.972 | IGHV1-18 | 3.3 | 5.57e-16 |
| IGHV3-21 | 2.5 | 3.00e-05 | IGHV3-74 | -1.7 | 0.001 | IGHV3-7 | -2.0 | 0.001 | IGHV1-69-2 | -0.6 | 0.990 | IGHV3-30 | 3.5 | 1.12e-15 |
| IGHV3-48 | 2.3 | 6.20e-05 | IGHV3-21 | -1.7 | 0.001 | IGHV4-4 | -1.4 | 0.002 | IGHV6-1 | -0.8 | 0.994 | IGHV1-46 | 2.7 | 6.46e-13 |
| IGHV3-20 | 2.1 | 9.71e-05 | IGHV5-10-1 | -1.1 | 0.003 | IGHV3-66 | -1.3 | 0.002 | IGHV3-73 | -0.8 | 0.996 | IGHV3-23 | 3.1 | 1.01e-12 |
| IGHV3-74 | 2.2 | 1.74e-04 | IGHV3-7 | -1.6 | 0.003 | IGHV3-73 | -1.2 | 0.003 | IGHV4-61 | -0.7 | 0.996 | IGHV3-74 | 2.7 | 1.32e-11 |
| IGHV2-70 | 1.8 | 4.67e-04 | IGHV3-11 | -1.4 | 0.004 | IGHV4-59 | -2.0 | 0.003 | IGHV3-11 | -0.9 | 0.996 | IGHV3-15 | 2.7 | 1.96e-11 |
| IGHV3-53 | 2.1 | 4.84e-04 | IGHV3-66 | -1.1 | 0.004 | IGHV5-10-1 | -1.2 | 0.003 | IGHV7-81 | -0.3 | 0.996 | IGHV3-33 | 2.9 | 3.27e-11 |
| IGHV3-66 | 1.9 | 8.59e-04 | IGHV1-18 | -1.5 | 0.004 | IGHV5-51 | -1.7 | 0.003 | IGHV2-5 | -0.8 | 0.996 | IGHV4-39 | 2.6 | 4.82e-11 |
| IGHV3-49 | 2.0 | 0.001 | IGHV3-20 | -1.2 | 0.004 | IGHV3-49 | -1.4 | 0.004 | IGHV3-64D | -0.5 | 0.996 | IGHV3-7 | 2.7 | 1.41e-10 |

| DM vs. NT |  |  | Mi2 vs. NT |  |  | NXP2 vs. NT |  |  | MDA5 vs. NT |  |  | TIF1γvs. NT |  |  |
| --- | --- | --- | --- | --- | --- | --- | --- | --- | --- | --- | --- | --- | --- | --- |
| Gene | LogFC | q-val | Gene | LogFC | q-val | Gene | LogFC | q-val | Gene | LogFC | q-val | Gene | LogFC | q-val |
| IGHV1-18 | 1.9 | 6.02e-05 | IGHV7-81 | -0.8 | 1.10e-05 | IGHV1-18 | 1.3 | 0.017 | IGHV5-51 | 3.4 | 5.79e-04 | IGHV3-53 | 1.9 | 2.18e-04 |
| IGHV3-7 | 2.1 | 1.94e-04 | IGHV3-38 | -0.8 | 3.64e-05 | IGHV3-64 | 1.1 | 0.036 | IGHV1-2 | 2.9 | 5.89e-04 | IGHV1-18 | 2.1 | 2.99e-04 |
| IGHV3-11 | 1.5 | 5.00e-04 | IGHV3-7 | 1.9 | 0.002 | IGHV3-7 | 1.4 | 0.049 | IGHV1-18 | 2.9 | 0.002 | IGHV3-7 | 2.5 | 4.92e-04 |
| IGHV3-53 | 1.3 | 0.001 | IGHV1-18 | 1.7 | 0.003 | IGHV4-39 | 1.1 | 0.072 | IGHV3-11 | 2.4 | 0.002 | IGHV4-59 | 2.1 | 0.002 |
| IGHV2-5 | 1.3 | 0.003 | IGHV4-34 | 1.5 | 0.004 | IGHV3-11 | 1.0 | 0.075 | IGHV3-7 | 2.9 | 0.003 | IGHV3-11 | 1.7 | 0.002 |
| IGHV4-34 | 1.3 | 0.004 | IGHV3-23 | 2.1 | 0.005 | IGHV7-81 | -0.5 | 0.102 | IGHV3-33 | 3.0 | 0.004 | IGHV3-23 | 2.6 | 0.002 |
| IGHV3-23 | 2.0 | 0.004 | IGHV1-69-2 | -0.9 | 0.006 | IGHV5-51 | 1.3 | 0.106 | IGHV4-59 | 2.8 | 0.004 | IGHV3-30 | 2.2 | 0.004 |
| IGHV4-59 | 1.8 | 0.004 | IGHV4-59 | 1.9 | 0.007 | IGHV2-5 | 1.0 | 0.111 | IGHV4-34 | 2.2 | 0.004 | IGHV1-69 | 1.4 | 0.007 |
| IGHV5-51 | 1.6 | 0.005 | IGHV3-74 | 1.4 | 0.007 | IGHV3-23 | 1.1 | 0.120 | IGHV2-5 | 2.6 | 0.006 | IGHV3-74 | 2.1 | 0.008 |
| IGHV3-30 | 1.9 | 0.007 | IGHV3-35 | -0.6 | 0.008 | IGHV4-34 | 0.9 | 0.127 | IGHV3-30 | 2.6 | 0.011 | IGHV1-69-2 | -0.8 | 0.011 |

| JO1 vs. NT |  |  | IMNM vs. NT |  |  | HMGCR vs. NT |  |  | SRP vs. NT |  |  | IBM vs. NT |  |  |
| --- | --- | --- | --- | --- | --- | --- | --- | --- | --- | --- | --- | --- | --- | --- |
| Gene | LogFC | q-val | Gene | LogFC | q-val | Gene | LogFC | q-val | Gene | LogFC | q-val | Gene | LogFC | q-val |
| IGHV3-7 | 4.7 | 1.81e-10 | IGHV1-18 | 1.2 | 0.008 | IGHV1-18 | 1.0 | 0.037 | IGHV4-59 | 2.4 | 2.37e-04 | IGHV1-18 | 5.1 | 1.92e-13 |
| IGHV1-18 | 4.0 | 1.21e-09 | IGHV3-7 | 1.2 | 0.029 | IGHV3-23 | 1.0 | 0.127 | IGHV3-7 | 2.3 | 3.55e-04 | IGHV1-46 | 3.8 | 4.21e-13 |
| IGHV3-20 | 3.0 | 1.22e-09 | IGHV3-23 | 1.3 | 0.044 | IGHV3-11 | 0.7 | 0.140 | IGHV3-23 | 2.3 | 0.002 | IGHV3-7 | 4.7 | 1.66e-12 |
| IGHV3-48 | 3.4 | 1.77e-09 | IGHV1-2 | 0.9 | 0.062 | IGHV3-30 | 1.0 | 0.142 | IGHV1-18 | 1.7 | 0.007 | IGHV3-43 | 3.0 | 1.94e-12 |
| IGHV3-21 | 3.9 | 6.18e-08 | IGHV3-48 | 0.8 | 0.084 | IGHV3-7 | 0.8 | 0.156 | IGHV1-2 | 1.7 | 0.008 | IGHV3-53 | 3.2 | 2.83e-12 |
| IGHV2-5 | 3.1 | 6.49e-08 | IGHV3-30 | 1.1 | 0.096 | IGHV2-5 | 0.6 | 0.199 | IGHV5-51 | 1.7 | 0.009 | IGHV4-34 | 3.5 | 4.27e-12 |
| IGHV1-46 | 2.8 | 1.87e-07 | IGHV3-11 | 0.7 | 0.099 | IGHV3-38 | -0.3 | 0.216 | IGHV2-70D | 1.2 | 0.009 | IGHV2-5 | 3.5 | 2.41e-11 |
| IGHV3-11 | 3.2 | 1.91e-07 | IGHV1-46 | 0.7 | 0.103 | IGHV7-81 | -0.3 | 0.221 | IGHV3-74 | 1.5 | 0.017 | IGHV3-30 | 5.3 | 4.45e-11 |
| IGHV3-43 | 2.6 | 2.02e-07 | IGHV4-59 | 1.0 | 0.105 | IGHV1-2 | 0.6 | 0.223 | IGHV3-43 | 1.1 | 0.030 | IGHV3-11 | 4.0 | 8.47e-11 |
| IGHV3-49 | 3.0 | 2.90e-07 | IGHV7-81 | -0.3 | 0.114 | IGHV3-48 | 0.6 | 0.237 | IGHV2-70 | 0.8 | 0.031 | IGHV3-48 | 3.4 | 9.50e-11 |

**Supplementary Table 4.** Expression of immunoglobulin heavy chain joining region genes across disease groups, shown as comparisons versus all other muscle biopsies (vs. ALL) and versus normal muscle biopsies (vs. NT). DM: dermatomyositis; IMNM: immune-mediated necrotizing myopathy; IBM: inclusion body myositis; NT: normal muscle biopsy.

| DM vs. ALL |  |  | Mi2 vs. ALL |  |  | NXP2 vs. ALL |  |  | MDA5 vs. ALL |  |  | TIF1γvs. ALL |  |  |
| --- | --- | --- | --- | --- | --- | --- | --- | --- | --- | --- | --- | --- | --- | --- |
| Gene | LogFC | q-val | Gene | LogFC | q-val | Gene | LogFC | q-val | Gene | LogFC | q-val | Gene | LogFC | q-val |
| IGHJ3 | -0.8 | 0.001 | IGHJ2 | -0.8 | 0.034 | IGHJ2 | -0.7 | 0.165 | IGHJ6 | -0.4 | 0.767 | IGHJ3 | -0.6 | 0.204 |
| IGHJ2 | -0.6 | 0.010 | IGHJ3 | -0.8 | 0.067 | IGHJ3 | -0.7 | 0.191 | IGHJ2 | 0.4 | 0.795 | IGHJ1 | -0.4 | 0.298 |
| IGHJ1 | -0.4 | 0.014 | IGHJ1 | -0.6 | 0.117 | IGHJ5 | -0.6 | 0.248 | IGHJ5 | 0.3 | 0.856 | IGHJ2 | -0.4 | 0.455 |
| IGHJ6 | -0.6 | 0.024 | IGHJ4 | -0.6 | 0.204 | IGHJ6 | -0.5 | 0.439 | IGHJ1 | 0.1 | 0.959 | IGHJ5 | -0.4 | 0.476 |
| IGHJ5 | -0.4 | 0.075 | IGHJ6 | -0.6 | 0.244 | IGHJ1 | -0.4 | 0.449 | IGHJ3 | -0.1 | 0.961 | IGHJ6 | -0.4 | 0.578 |
| IGHJ4 | -0.4 | 0.087 | IGHJ5 | -0.4 | 0.395 | IGHJ4 | -0.4 | 0.586 | IGHJ4 | 0.1 | 0.976 | IGHJ4 | -0.3 | 0.626 |

| JO1 vs. ALL |  |  | IMNM vs. ALL |  |  | HMGCR vs. ALL |  |  | SRP vs. ALL |  |  | IBM vs. ALL |  |  |
| --- | --- | --- | --- | --- | --- | --- | --- | --- | --- | --- | --- | --- | --- | --- |
| Gene | LogFC | q-val | Gene | LogFC | q-val | Gene | LogFC | q-val | Gene | LogFC | q-val | Gene | LogFC | q-val |
| IGHJ5 | 1.0 | 0.053 | IGHJ4 | -0.5 | 0.040 | IGHJ4 | -0.6 | 0.050 | IGHJ6 | -0.6 | 0.991 | IGHJ2 | 1.0 | 1.16e-04 |
| IGHJ3 | 0.9 | 0.057 | IGHJ2 | -0.5 | 0.064 | IGHJ5 | -0.5 | 0.171 | IGHJ2 | -0.4 | 0.996 | IGHJ6 | 1.2 | 3.96e-04 |
| IGHJ2 | 0.7 | 0.202 | IGHJ6 | -0.5 | 0.108 | IGHJ2 | -0.4 | 0.178 | IGHJ1 | -0.3 | 0.996 | IGHJ4 | 1.0 | 0.001 |
| IGHJ4 | 0.7 | 0.216 | IGHJ1 | -0.3 | 0.123 | IGHJ1 | -0.3 | 0.248 | IGHJ4 | -0.2 | 0.996 | IGHJ1 | 0.5 | 0.020 |
| IGHJ1 | 0.5 | 0.238 | IGHJ5 | -0.4 | 0.158 | IGHJ6 | -0.4 | 0.368 | IGHJ5 | -0.1 | 0.996 | IGHJ3 | 0.7 | 0.036 |
| IGHJ6 | 0.5 | 0.529 | IGHJ3 | -0.2 | 0.563 | IGHJ3 | -0.3 | 0.444 | IGHJ3 | 0.1 | 0.996 | IGHJ5 | 0.6 | 0.064 |

| DM vs. NT |  |  | Mi2 vs. NT |  |  | NXP2 vs. NT |  |  | MDA5 vs. NT |  |  | TIF1γvs. NT |  |  |
| --- | --- | --- | --- | --- | --- | --- | --- | --- | --- | --- | --- | --- | --- | --- |
| Gene | LogFC | q-val | Gene | LogFC | q-val | Gene | LogFC | q-val | Gene | LogFC | q-val | Gene | LogFC | q-val |
| IGHJ1 | -0.3 | 0.145 | IGHJ1 | -0.5 | 0.029 | IGHJ3 | -0.5 | 0.213 | IGHJ2 | 0.6 | 0.167 | IGHJ1 | -0.4 | 0.175 |
| IGHJ3 | -0.4 | 0.196 | IGHJ2 | -0.5 | 0.034 | IGHJ1 | -0.4 | 0.233 | IGHJ5 | 0.5 | 0.355 | IGHJ3 | -0.4 | 0.291 |
| IGHJ2 | -0.1 | 0.696 | IGHJ3 | -0.6 | 0.048 | IGHJ2 | -0.4 | 0.278 | IGHJ4 | 0.4 | 0.516 | IGHJ4 | 0.2 | 0.638 |
| IGHJ6 | -0.1 | 0.697 | IGHJ4 | -0.2 | 0.443 | IGHJ5 | -0.4 | 0.347 | IGHJ6 | -0.2 | 0.714 | IGHJ5 | -0.1 | 0.889 |
| IGHJ4 | 0.0 | 0.887 | IGHJ6 | -0.2 | 0.528 | IGHJ6 | -0.2 | 0.645 | IGHJ3 | 0.1 | 0.899 | IGHJ2 | 0.0 | 0.942 |
| IGHJ5 | 0.0 | 0.901 | IGHJ5 | -0.2 | 0.609 | IGHJ4 | 0.0 | 0.956 | IGHJ1 | 0.0 | 0.953 | IGHJ6 | 0.0 | 0.974 |

| JO1 vs. NT |  |  | IMNM vs. NT |  |  | HMGCR vs. NT |  |  | SRP vs. NT |  |  | IBM vs. NT |  |  |
| --- | --- | --- | --- | --- | --- | --- | --- | --- | --- | --- | --- | --- | --- | --- |
| Gene | LogFC | q-val | Gene | LogFC | q-val | Gene | LogFC | q-val | Gene | LogFC | q-val | Gene | LogFC | q-val |
| IGHJ5 | 1.0 | 0.001 | IGHJ1 | -0.3 | 0.163 | IGHJ1 | -0.3 | 0.224 | IGHJ3 | 0.5 | 0.207 | IGHJ2 | 1.2 | 5.96e-06 |
| IGHJ2 | 0.8 | 0.002 | IGHJ6 | -0.1 | 0.802 | IGHJ4 | -0.2 | 0.620 | IGHJ1 | -0.2 | 0.345 | IGHJ4 | 1.2 | 4.87e-05 |
| IGHJ4 | 0.9 | 0.003 | IGHJ4 | -0.1 | 0.802 | IGHJ5 | -0.1 | 0.743 | IGHJ6 | -0.2 | 0.538 | IGHJ6 | 1.3 | 4.24e-04 |
| IGHJ3 | 1.0 | 0.017 | IGHJ2 | -0.1 | 0.822 | IGHJ2 | -0.1 | 0.846 | IGHJ4 | 0.2 | 0.599 | IGHJ3 | 0.8 | 0.005 |
| IGHJ6 | 0.7 | 0.052 | IGHJ3 | 0.0 | 0.906 | IGHJ3 | -0.1 | 0.869 | IGHJ5 | 0.2 | 0.618 | IGHJ5 | 0.8 | 0.008 |
| IGHJ1 | 0.4 | 0.127 | IGHJ5 | 0.0 | 0.907 | IGHJ6 | 0.0 | 0.973 | IGHJ2 | 0.1 | 0.848 | IGHJ1 | 0.5 | 0.049 |

**Supplementary Table 5.**
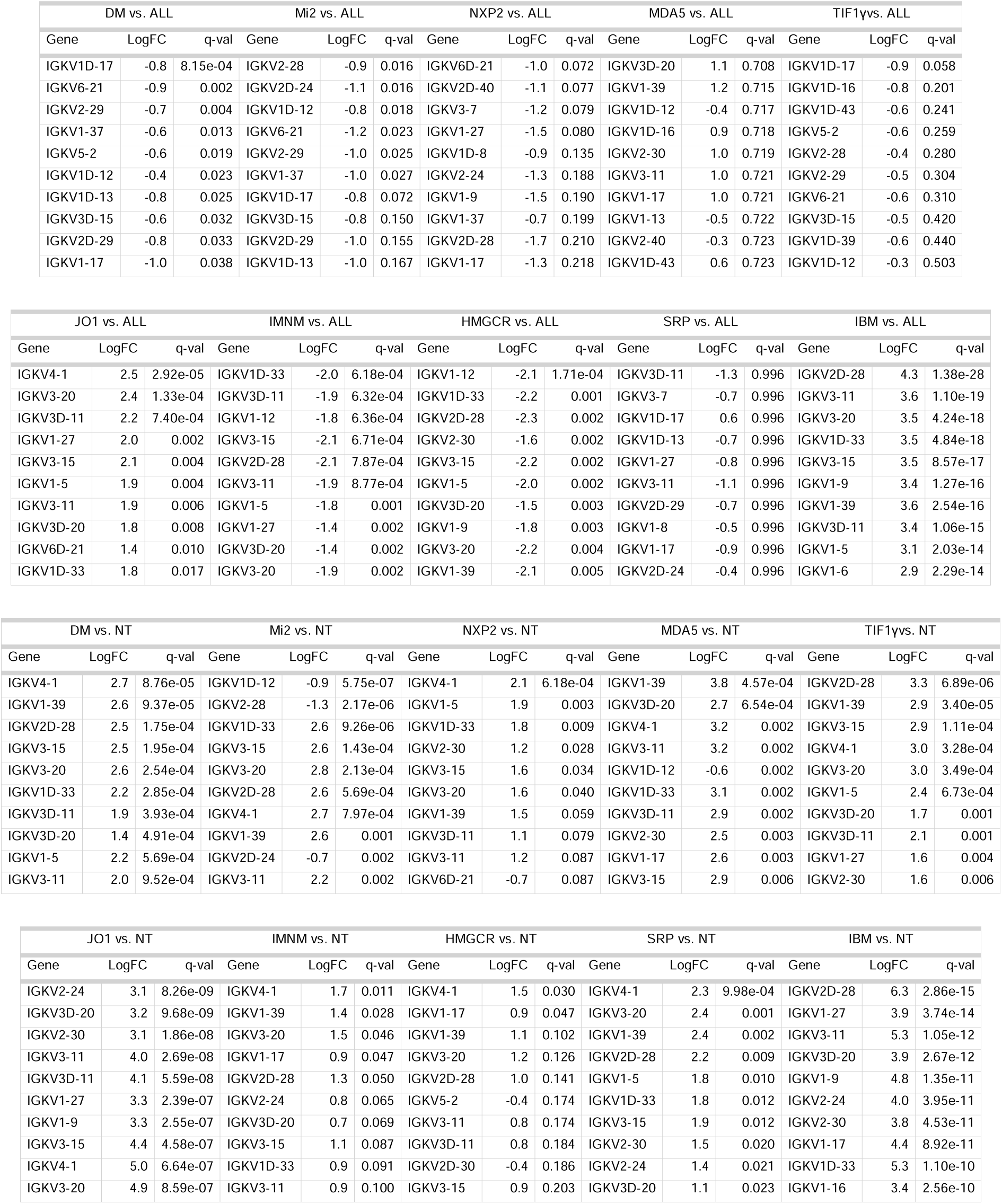
Expression of top 10 immunoglobulin kappa variable region genes across disease groups, shown as comparisons versus all other muscle biopsies (vs. ALL) and versus normal muscle biopsies (vs. NT). DM: dermatomyositis; IMNM: immune-mediated necrotizing myopathy; IBM: inclusion body myositis; NT: normal muscle biopsy.

**Supplementary Table 6.** Expression of top 10 immunoglobulin lambda variable region genes across disease groups, shown as comparisons versus all other muscle biopsies (vs. ALL) and versus normal muscle biopsies (vs. NT). DM: dermatomyositis; IMNM: immune-mediated necrotizing myopathy; IBM: inclusion body myositis; NT: normal muscle biopsy.

| DM vs. ALL |  |  | Mi2 vs. ALL |  |  | NXP2 vs. ALL |  |  | MDA5 vs. ALL |  |  | TIF1vs. ALL |  |  |
| --- | --- | --- | --- | --- | --- | --- | --- | --- | --- | --- | --- | --- | --- | --- |
| Gene | LogFC | q-val | Gene | LogFC | q-val | Gene | LogFC | q-val | Gene | LogFC | q-val | Gene | LogFC | q-val |
| IGLV4-3 | -0.6 | 2.13e-04 | IGLV5-37 | -0.9 | 0.012 | IGLV3-9 | -1.5 | 0.068 | IGLV3-21 | 2.0 | 0.406 | IGLV5-37 | -0.8 | 0.056 |
| IGLV5-37 | -0.6 | 9.71e-04 | IGLV3-32 | -0.7 | 0.017 | IGLV5-52 | 1.2 | 0.116 | IGLV7-43 | 1.3 | 0.614 | IGLV4-3 | -0.6 | 0.075 |
| IGLV3-32 | -0.5 | 9.71e-04 | IGLV4-3 | -0.7 | 0.025 | IGLV3-1 | -1.3 | 0.254 | IGLV2-11 | 1.5 | 0.632 | IGLV10-54 | -0.9 | 0.107 |
| IGLV5-48 | -0.6 | 0.001 | IGLV2-33 | -0.8 | 0.040 | IGLV4-69 | -1.0 | 0.279 | IGLV1-51 | 1.6 | 0.667 | IGLV3-22 | -0.5 | 0.127 |
| IGLV2-33 | -0.6 | 0.003 | IGLV3-22 | -0.6 | 0.048 | IGLV3-25 | -1.2 | 0.295 | IGLV3-16 | 0.9 | 0.670 | IGLV5-48 | -0.6 | 0.131 |
| IGLV3-22 | -0.4 | 0.005 | IGLV7-43 | -1.1 | 0.073 | IGLV2-8 | -1.3 | 0.336 | IGLV6-57 | 1.1 | 0.692 | IGLV5-52 | -1.0 | 0.147 |
| IGLV10-54 | -0.7 | 0.015 | IGLV5-48 | -0.6 | 0.073 | IGLV1-44 | -1.2 | 0.356 | IGLV1-47 | 1.1 | 0.701 | IGLV3-32 | -0.4 | 0.156 |
| IGLV3-16 | -0.4 | 0.050 | IGLV3-16 | -0.7 | 0.089 | IGLV10-54 | -0.6 | 0.377 | IGLV3-27 | 0.8 | 0.705 | IGLV8-61 | -1.0 | 0.182 |
| IGLV7-43 | -0.6 | 0.072 | IGLV3-12 | -0.7 | 0.110 | IGLV5-48 | -0.4 | 0.384 | IGLV3-10 | 1.1 | 0.706 | IGLV3-16 | -0.6 | 0.194 |
| IGLV4-60 | -0.5 | 0.081 | IGLV4-60 | -0.7 | 0.224 | IGLV4-60 | -0.6 | 0.390 | IGLV1-44 | 1.1 | 0.708 | IGLV7-43 | -0.8 | 0.220 |

| JO1 vs. ALL |  |  | IMNM vs. ALL |  |  | HMGR vs. ALL |  |  | SRP vs. ALL |  |  | IBM vs. ALL |  |  |
| --- | --- | --- | --- | --- | --- | --- | --- | --- | --- | --- | --- | --- | --- | --- |
| Gene | LogFC | q-val | Gene | LogFC | q-val | Gene | LogFC | q-val | Gene | LogFC | q-val | Gene | LogFC | q-val |
| IGLV3-25 | 2.3 | 6.68e-05 | IGLV3-21 | -2.2 | 6.82e-05 | IGLV3-21 | -2.3 | 1.82e-04 | IGLV3-10 | -1.2 | 0.811 | IGLV2-23 | 3.7 | 1.68e-19 |
| IGLV1-44 | 2.0 | 0.001 | IGLV1-40 | -2.2 | 9.20e-05 | IGLV3-19 | -2.1 | 2.80e-04 | IGLV1-51 | -1.3 | 0.993 | IGLV2-8 | 3.4 | 2.74e-17 |
| IGLV1-40 | 2.1 | 0.002 | IGLV3-10 | -1.5 | 9.75e-05 | IGLV3-25 | -1.9 | 3.33e-04 | IGLV3-16 | -0.5 | 0.996 | IGLV1-40 | 3.4 | 7.54e-16 |
| IGLV3-19 | 2.0 | 0.002 | IGLV3-19 | -1.9 | 1.33e-04 | IGLV1-40 | -2.4 | 3.89e-04 | IGLV1-50 | -0.4 | 0.996 | IGLV1-51 | 3.6 | 4.03e-15 |
| IGLV3-1 | 2.0 | 0.002 | IGLV3-25 | -1.6 | 7.48e-04 | IGLV2-11 | -2.1 | 8.41e-04 | IGLV3-27 | -0.5 | 0.996 | IGLV1-44 | 2.9 | 1.30e-12 |
| IGLV6-57 | 1.8 | 0.003 | IGLV6-57 | -1.5 | 0.001 | IGLV2-23 | -2.3 | 0.001 | IGLV5-37 | -0.3 | 0.996 | IGLV2-14 | 2.9 | 1.54e-11 |
| IGLV2-8 | 2.0 | 0.003 | IGLV1-51 | -1.8 | 0.001 | IGLV6-57 | -1.7 | 0.001 | IGLV1-40 | -1.0 | 0.996 | IGLV3-1 | 2.8 | 1.84e-11 |
| IGLV2-11 | 1.9 | 0.006 | IGLV2-23 | -1.9 | 0.002 | IGLV1-44 | -2.1 | 0.001 | IGLV9-49 | -0.4 | 0.996 | IGLV3-19 | 2.8 | 3.55e-11 |
| IGLV2-23 | 1.9 | 0.007 | IGLV2-11 | -1.7 | 0.002 | IGLV2-8 | -2.0 | 0.003 | IGLV8-61 | -0.6 | 0.996 | IGLV3-9 | 2.6 | 3.42e-10 |
| IGLV4-60 | 1.4 | 0.009 | IGLV2-8 | -1.8 | 0.002 | IGLV3-9 | -1.4 | 0.003 | IGLV3-19 | -0.8 | 0.996 | IGLV6-57 | 2.4 | 4.10e-10 |

| DM vs. NT |  |  | Mi2 vs. NT |  |  | NXP2 vs. NT |  |  | MDA5 vs. NT |  |  | TIF1vs. NT |  |  |
| --- | --- | --- | --- | --- | --- | --- | --- | --- | --- | --- | --- | --- | --- | --- |
| Gene | LogFC | q-val | Gene | LogFC | q-val | Gene | LogFC | q-val | Gene | LogFC | q-val | Gene | LogFC | q-val |
| IGLV6-57 | 2.4 | 1.45e-07 | IGLV1-40 | 3.4 | 3.46e-07 | IGLV6-57 | 1.8 | 1.11e-04 | IGLV6-57 | 3.3 | 1.01e-04 | IGLV6-57 | 2.7 | 1.60e-06 |
| IGLV1-40 | 2.9 | 6.71e-07 | IGLV3-32 | -0.9 | 5.75e-07 | IGLV2-23 | 2.4 | 4.58e-04 | IGLV3-21 | 4.0 | 5.12e-04 | IGLV1-40 | 3.0 | 3.34e-06 |
| IGLV2-23 | 2.8 | 1.96e-05 | IGLV5-37 | -1.1 | 4.85e-06 | IGLV3-21 | 2.0 | 0.003 | IGLV2-11 | 3.5 | 5.67e-04 | IGLV1-44 | 2.8 | 3.35e-05 |
| IGLV1-44 | 2.6 | 2.15e-05 | IGLV4-3 | -0.8 | 8.83e-06 | IGLV1-40 | 2.0 | 0.004 | IGLV1-51 | 3.4 | 9.35e-04 | IGLV3-25 | 2.2 | 6.12e-05 |
| IGLV3-21 | 2.5 | 9.46e-05 | IGLV3-22 | -0.8 | 1.44e-05 | IGLV2-14 | 2.2 | 0.009 | IGLV1-40 | 3.6 | 0.001 | IGLV2-23 | 2.9 | 1.71e-04 |
| IGLV3-25 | 1.8 | 2.09e-04 | IGLV2-23 | 3.0 | 6.75e-05 | IGLV2-11 | 1.8 | 0.009 | IGLV1-44 | 3.6 | 0.002 | IGLV5-37 | -0.9 | 2.88e-04 |
| IGLV2-11 | 2.0 | 8.14e-04 | IGLV1-44 | 2.6 | 1.83e-04 | IGLV1-44 | 1.5 | 0.020 | IGLV1-47 | 2.8 | 0.002 | IGLV4-69 | 1.9 | 3.24e-04 |
| IGLV3-32 | -0.6 | 8.14e-04 | IGLV2-8 | 2.6 | 4.53e-04 | IGLV3-19 | 1.5 | 0.033 | IGLV2-8 | 2.9 | 0.003 | IGLV3-1 | 2.3 | 3.27e-04 |
| IGLV4-3 | -0.6 | 8.66e-04 | IGLV6-57 | 2.0 | 4.81e-04 | IGLV1-47 | 1.1 | 0.064 | IGLV3-10 | 2.2 | 0.007 | IGLV3-21 | 2.6 | 3.28e-04 |
| IGLV3-1 | 1.9 | 9.59e-04 | IGLV3-25 | 2.0 | 6.43e-04 | IGLV5-48 | -0.6 | 0.081 | IGLV3-1 | 2.7 | 0.007 | IGLV4-3 | -0.7 | 3.28e-04 |

| JO1 vs. NT |  |  | IMNM vs. NT |  |  | HMGR vs. NT |  |  | SRP vs. NT |  |  | IBM vs. NT |  |  |
| --- | --- | --- | --- | --- | --- | --- | --- | --- | --- | --- | --- | --- | --- | --- |
| Gene | LogFC | q-val | Gene | LogFC | q-val | Gene | LogFC | q-val | Gene | LogFC | q-val | Gene | LogFC | q-val |
| IGLV3-25 | 3.9 | 4.73e-11 | IGLV6-57 | 1.2 | 0.002 | IGLV6-57 | 1.0 | 0.019 | IGLV1-44 | 2.7 | 3.09e-05 | IGLV6-57 | 4.4 | 4.38e-18 |
| IGLV6-57 | 3.9 | 2.48e-10 | IGLV2-23 | 1.5 | 0.008 | IGLV5-52 | -1.3 | 0.028 | IGLV2-23 | 2.7 | 5.64e-05 | IGLV1-51 | 5.1 | 1.89e-13 |
| IGLV1-40 | 4.6 | 1.21e-08 | IGLV5-52 | -1.4 | 0.012 | IGLV5-48 | -0.5 | 0.040 | IGLV2-14 | 2.4 | 7.48e-04 | IGLV3-1 | 4.5 | 2.47e-13 |
| IGLV2-23 | 4.6 | 1.61e-08 | IGLV1-44 | 1.4 | 0.014 | IGLV2-23 | 1.1 | 0.066 | IGLV6-57 | 1.8 | 8.63e-04 | IGLV3-10 | 3.3 | 4.45e-13 |
| IGLV3-1 | 3.8 | 5.05e-08 | IGLV5-48 | -0.5 | 0.024 | IGLV1-44 | 1.0 | 0.107 | IGLV3-1 | 2.0 | 0.001 | IGLV2-23 | 6.0 | 8.21e-13 |
| IGLV1-44 | 4.3 | 5.55e-08 | IGLV1-40 | 1.1 | 0.031 | IGLV1-40 | 0.9 | 0.112 | IGLV2-11 | 2.0 | 0.002 | IGLV2-8 | 5.2 | 2.32e-12 |
| IGLV2-14 | 4.0 | 4.61e-07 | IGLV2-14 | 1.2 | 0.055 | IGLV3-32 | -0.4 | 0.112 | IGLV1-40 | 1.8 | 0.008 | IGLV1-44 | 5.1 | 2.54e-11 |
| IGLV2-8 | 4.1 | 4.62e-07 | IGLV5-37 | -0.5 | 0.056 | IGLV3-22 | -0.4 | 0.112 | IGLV2-8 | 1.9 | 0.008 | IGLV1-40 | 5.6 | 3.14e-11 |
| IGLV1-51 | 3.6 | 8.26e-07 | IGLV1-47 | 0.9 | 0.057 | IGLV1-47 | 0.8 | 0.121 | IGLV3-25 | 1.6 | 0.016 | IGLV2-18 | 2.8 | 5.25e-11 |
| IGLV2-11 | 3.8 | 1.16e-06 | IGLV3-32 | -0.4 | 0.059 | IGLV5-37 | -0.4 | 0.134 | IGLV3-21 | 1.4 | 0.039 | IGLV3-25 | 4.1 | 6.21e-11 |

